# Single-Nucleus to Whole Body Phenotyping Reveals Neuromuscular Impairment and Preserved Exercise Adaptations in Long-Term Pediatric HSCT Survivors >10 years after treatment

**DOI:** 10.64898/2026.04.24.26351644

**Authors:** Casper Soendenbroe, Anne Nissen, Lise Marie Krogh, Peter Schjerling, Jasmin Garoussian, Veronika D. Storm, Michael Kjaer, Jesper L. Andersen, Kenneth H. Mertz, Martin Kaj Fridh, Klaus Müller, Abigail L. Mackey

## Abstract

Allogeneic hematopoietic stem cell transplantation (HSCT) is a life-saving treatment for hematologic malignancies, but long-term survivors present with lower muscle mass and functional capacity. In adult HSCT survivors 10-20 years after treatment, single nucleus RNA sequencing uncovered elevated XRRA1 expression levels in all muscle nuclei populations, which was retained in primary muscle stem cell cultures. HSCT survivors were characterized *in vivo* by impaired neuromuscular innervation that associated with muscle weakness, and lower muscle stem cell neurotrophic action. Despite these impairments, the molecular and physiological responses to heavy resistance training (HReT) were preserved in HSCT survivors, as demonstrated in a pre-registered clinical trial (ClinicalTrials.gov: NCT04922970). After 12 weeks of HReT, gains in muscle mass and strength were similar in HSCT survivors and healthy controls. In addition, we observed that ∼9% of muscle-resident immune cells persist into adulthood and that bone marrow derived cells do not adopt alternative cell fates in muscle tissue, resolving long-standing questions in human muscle biology. Together, these findings uncover molecular mechanisms of HSCT sequelae in muscle nuclei and muscle stem cells, which, importantly, can at least partly be overcome by mechanical loading. Given the growing population of HSCT survivors and the multitude of benefits of HReT for all organ systems, our findings support the importance of HReT in this population to promote healthspan.

**GRAPHICAL ABSTRACT:** 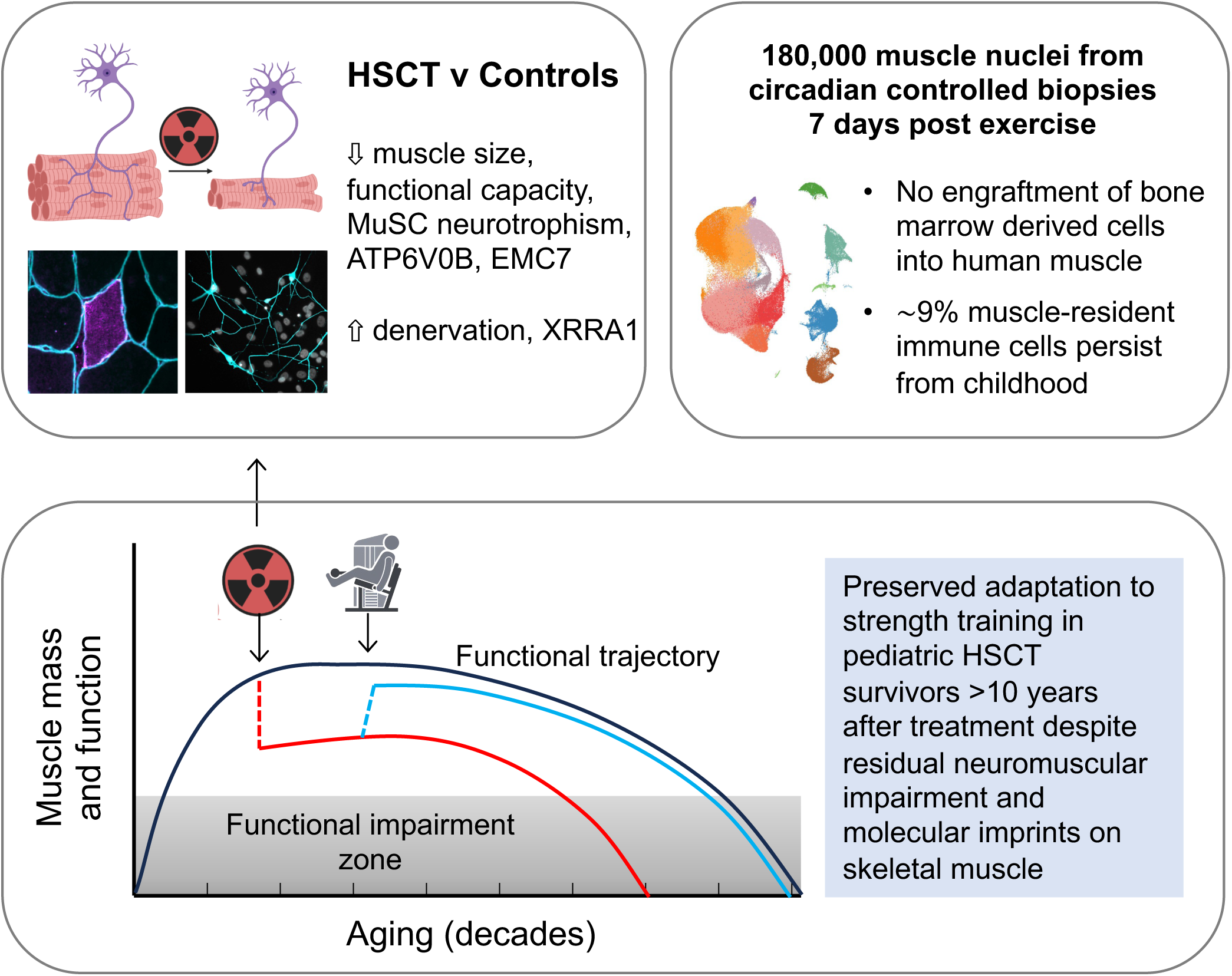

## INTRODUCTION

Pediatric allogeneic hematopoietic stem cell transplantation (HSCT) is a life-saving treatment for malignant and benign hematopoietic diseases^1^, with substantial improvement in survival rates in recent decades. Recipients who survive at least the first two years from transplant now have survival rates approaching general population levels^2^. As more pediatric HSCT recipients survive into adulthood, and the first adult survivors are now reaching mid-life, clinical attention has increasingly shifted toward managing long-term sequelae following pediatric HSCT^3,4^. Among these, reductions in skeletal muscle mass and function, as well as neuromuscular complications, are well-documented, with little evidence of spontaneous recovery even years after treatment^5–9^. While exercise interventions in this population are limited, findings regarding the feasibility and potential benefits of exercise interventions initiated before, during, or shortly after pediatric HSCT^10–13^, are generally encouraging. However, the only study to evaluate muscle adaptations in survivors of pediatric HSCT more than one year post treatment (mean follow-up of 8.4 years) found that six months of combined resistance and aerobic exercise failed to increase muscle mass^14^. While this may suggest impaired muscle plasticity, the absence of a healthy control group limits this interpretation. Moreover, we lack basic mechanistic insight into the molecular underpinnings of the persistent muscle dysfunction in long-term HSCT survivors and how this may limit the potential of exercise to restore muscle mass and function in this population.

Both age- and disease-related loss of muscle mass and function are key risk factors for disability and mortality^15–17^, underlining the importance of targeted strategies to restore and maintain skeletal muscle throughout the lifespan. Mechanical loading is the most potent stimulus for increasing skeletal muscle mass and function, and the cellular interplay driving this adaptation is largely understood^18,19^. The challenge of maintaining the transcriptional demand of the growing multinucleated muscle fiber is met by addition of myonuclei, derived from resident muscle stem cells^20^. In concert with the other resident cell populations of the muscle, such as fibroblasts, immune cells and vascular cells, muscle stem cells work to maintain, repair and facilitate growth of the muscle fibers^21^. However, an unsolved question in muscle biology relates to the relative contribution of tissue-resident versus bone marrow-derived immune cells to the overall immune cell landscape of skeletal muscle. While resident tissue macrophages originate from embryonic development and persist into adulthood independently of circulating monocytes for some tissues^22^, bone marrow-derived leukocytes home to damaged muscle in healthy young individuals^23^, demonstrating a role for both populations. Interestingly, rodent studies using labelled bone marrow-derived cells indicate incorporation into non-hematopoietic tissues, including muscle fibers themselves^24–27^. In humans, *in situ* hybridization for Y-chromosome from male donors demonstrated rare occurrences of bone marrow-derived cells in female HSCT recipient tissues^28,29^, but with the advent of snRNAseq, this question can be addressed more robustly. Ablation of the embryonically derived resident muscle macrophage population by chemotherapy and radiation (part of the HSCT conditioning regimen) could increase dependence on bone marrow-derived cells and may have implications for the growth and adaptive potential of skeletal muscle. Pediatric HSCT survivors go through many years of childhood and adolescent skeletal growth after their treatment, and it is unknown how the HSCT treatment regimen impacts the various muscle cell types during growth and in response to exercise. This gap in knowledge limits the development of targeted interventions to mitigate, and potentially restore, post-HSCT musculoskeletal deficits.

We hypothesized that survivors of pediatric HSCT would exhibit cellular and molecular defects underlying impaired muscle mass and function, accompanied by altered acute responses and reduced adaptive capacity to heavy resistance exercise training (HReT), compared with healthy controls. To this end, we conducted the first in-depth investigation of skeletal muscle of long-term pediatric HSCT survivors before and after a single bout, or 12 weeks, of HReT, by comprehensively characterizing structural, cellular, and transcriptional features of skeletal muscle and peripheral blood mononuclear cells, in parallel with extensive *in vivo* phenotyping and testing.

## RESULTS

### Description of the study and participants

To characterize the physiological profile of pediatric hematopoietic stem cell transplantation (HSCT) survivors, we enrolled 12 female and 6 male long-term survivors through the outpatient clinic at the National Center of Pediatric HSCT, Copenhagen University Hospital – Rigshospitalet, Copenhagen, Denmark. Eighteen female and 10 male age-matched controls were recruited for comparison. The HSCT and control groups were similar for sex distribution, age, and BMI; however, height and weight were modestly lower in the HSCT group (both p < 0.05) (Table S1). Disease and transplant characteristics of all pediatric HSCT participants, including those enrolled in the single bout and 12-week training, are summarized in Table S2. Briefly, the median [range] age at transplantation was 11 [1–17] years, and the time since HSCT at study enrollment was 16 [10–29] years. Most HSCT survivors (89%) had a malignant underlying disease, all received chemotherapy as part of their preparative conditioning therapy before the transplantation, and 56% additionally underwent total body irradiation therapy. All patients experienced acute graft-versus-host disease, with 50% developing moderate to severe manifestations (grade 2–4).

Participants underwent extensive baseline phenotyping and subsequently completed either a single bout or a 12-week program of HReT. Assessment of health-related quality of life using the SF-36 questionnaire revealed a lower General Health domain score in HSCT survivors compared with controls (Mann–Whitney U test, p = 0.035; median [range]: HSCT, 60 [25–100] vs control, 80 [60–100]; Supplemental Data 1), whereas no significant differences were observed for the remaining SF-36 domains (controls: n = 17; HSCT: n = 11). Resting systolic (SBP) and diastolic (DBP) blood pressure, mean arterial pressure (MAP), and pulse pressure (PP) did not differ between groups and were within normal reference ranges in both cohorts (controls, n = 17; HSCT, n = 11; unpaired t-test; SBP: HSCT 111 ± 12 vs control 115 ± 13 mmHg; DBP: HSCT 72 ± 9 vs control 74 ± 9 mmHg; MAP: HSCT 85 ± 9 vs control 89 ± 10 mmHg; PP: HSCT 40 ± 9 vs control 44 ± 8; Supplemental Data 1). Blood biochemistry profiling demonstrated modest group differences in selected hematological, endocrine, and nutritional markers, including leukocyte count, monocytes, immature granulocytes, triiodothyronine (T3), alkaline phosphatase, ferritin, and 25-hydroxyvitamin D (all p < 0.05), whereas the majority of routine hematological, inflammatory, metabolic, renal, hepatic, and lipid parameters were comparable between groups (Table S3). To further evaluate systemic metabolic health, participants underwent a 3-hour oral glucose tolerance test (OGTT; Figure S1). While plasma glucose area under the curve (AUC) did not differ between groups, HSCT survivors exhibited a higher C-peptide AUC compared with controls (controls: n = 17; HSCT: n = 9; p < 0.05), indicative of reduced whole-body insulin sensitivity.

### Reduced muscle mass and strength in pediatric HSCT survivors

Magnetic resonance imaging (MRI) and dual-energy X-ray absorptiometry (DXA) revealed that, compared with healthy controls, HSCT survivors exhibited 18 % smaller quadriceps muscle cross-sectional area (p < 0.05), and 16 % and 14 % lower leg and whole-body lean mass, respectively (both p < 0.05; Figure 1A, B). Whole-body bone mineral density (p = 0.0614) and Z-score (p = 0.0667) tended to be lower in HSCT survivors compared with controls (Figure 1C). Maximal voluntary contractile strength (MVC) was 25 % lower in HSCT survivors (p < 0.01; 17% lower when normalization to body weight), and rate of force development (RFD) was 29 % lower in HSCT survivors (p < 0.001; Figure 1D). Functional performance assessed by the 30-s chair-stand test and the 6-min walk test did not differ between groups, whereas performance in the timed up-and-go test was significantly impaired in HSCT survivors compared with controls (p < 0.05; Figure 1E). Collectively, these findings confirm a markedly reduced musculoskeletal profile in long-term HSCT survivors, consistent with prior observations^7^.

**Figure 1.**
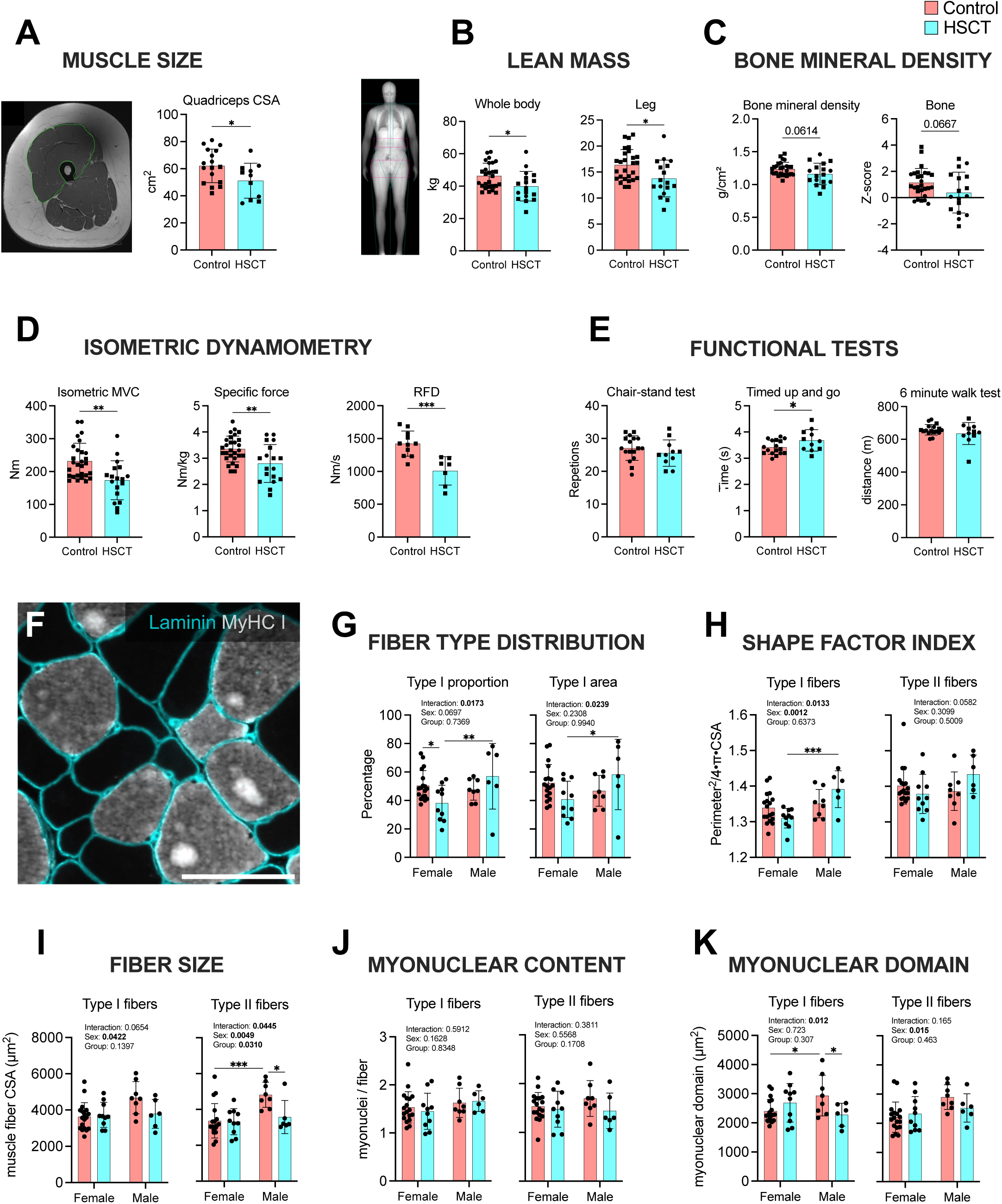
Reduced muscle mass, strength, and fiber size in long-term HSCT survivors. (A) Representative magnetic resonance imaging (MRI) of the thigh and quadriceps muscle cross-sectional area (CSA) assessed by MRI (controls: n = 17; HSCT: n = 11). (B) Representative DXA scan and whole-body, and leg, lean mass assessed by DXA (controls: n = 27; HSCT: n = 17). (C) Whole-body bone mineral density and Z-scores assessed by DXA (controls: n = 27; HSCT: n = 17). (D) Isometric maximal voluntary contraction (MVC) measured by dynamometry, presented as Isometric MVC and Specific force (normalized to body weight) (controls: n = 28; HSCT: n = 18). Rate of force development (RFD) was also measured by dynamometry (controls: n = 11; HSCT: n = 7). (E) Physical function assessed using the 30-s chair-stand test, timed up-and-go test, and 6-min walk test (controls: n = 17; HSCT: n = 11). F-K. Data from muscle biopsy immunofluorescence, analyzed by two-way repeated-measures ANOVA with factors Group and Sex. Data are displayed as individual values with mean ± SD. Men are represented by circles, women by squares (controls: n = 26; HSCT: n = 16). (F) Muscle biopsy cross-section stained by immunofluorescence with the basement membrane marker laminin, delineating the muscle fiber borders, and type I myosin heavy chain (MyHC I). The unstained (black) fibers were designated type II fibers. Scale bar = 100 μm. Such images were used to calculate fiber type-specific data for G) fiber type distribution (proportion and relative area), H) shape factor index, I) fiber cross-sectional area (CSA), J) myonuclear content, K) myonuclear domain size. A-E. Data were analyzed with unpaired two-tailed t-tests.

Immunofluorescence analysis of biopsy cryosections (Figure 1F) revealed an influence of both group (HSCT vs. control) and sex on fiber type proportions (Figure 1G). Female HSCT survivors had a lower proportion of type I fibers compared with female controls (p < 0.05), which was not the case in male HSCT survivors, although we cannot rule out selection bias in the healthy controls that volunteered for this study. To account for these fiber type differences, all additional microscopy data are presented in fiber type specific manner. Fiber morphology was characterized using the Shape Factor Index (SFI)^30,31^, which showed greater type I fiber shape abnormalities in male HSCT survivors compared with male controls, while female HSCT participants were largely comparable to female controls (Figure 1H). In accordance with the reduced muscle mass and strength at the whole-body level, type II myofiber cross-sectional area (CSA) was significantly smaller in HSCT survivors compared with controls (Figure 1I), a difference driven by the male participants (p < 0.05). Female HSCT survivors displayed similar type I and type II fiber CSA to their healthy counterparts.

To gain more insight into factors regulating myofiber size, we next examined potential group differences in the number of myonuclei. While we found no differences in the number of myonuclei per fiber (Figure 1J), a significant group × sex interaction was observed for type I fiber myonuclear domain (Figure 1K). Post hoc testing revealed smaller domains in male HSCT survivors compared with male controls (p < 0.05), whereas the opposite pattern was evident in females. In addition, a main effect of sex was observed for type II fiber myonuclear domain. These findings suggest that disease and/or treatment history may influence myonuclear regulation in a sex-specific manner, potentially affecting fiber maintenance or transcriptional capacity.

### Single nucleus RNA sequencing of muscle tissue and circulating immune cells

To compare the transcriptional profiles of circulating immune cells, and muscle cells, between HSCT survivors and healthy controls, we performed single-nucleus RNA sequencing (snRNAseq) on isolated peripheral blood mononuclear cells (PBMC) alongside skeletal muscle biopsy tissue from a subset of participants (n = 4 HSCT survivors and n = 4 controls). Muscle biopsies were collected from both vastus lateralis muscles from each participant 7 days after a unilateral bout of HReT, giving rise to 16 muscle tissue samples and 8 PBMC samples in total. Participants were selected based on the availability of sufficient biopsy material, and participant characteristics are summarized in Table S1B. RNA in isolated nuclei from PBMCs and muscle tissue were sequenced on the 10x Genomics Chromium Next GEM Single Cell 3’ v3.1 platform. After quality control filtering to remove doublets and low-quality nuclei, a total of 118,271 PBMC nuclei remained, corresponding to 14,784 ± 5,664 (mean ± SD) nuclei per participant (8 participants). Correspondingly for the muscle, 180,419 nuclei were retained, with an average of 11,276 ± 3,804 nuclei per tissue sample (22,552 ± 7,392 nuclei per subject) (Supplemental Data 2).

### Peripheral immune landscapes are largely indistinguishable between HSCT survivors and controls

Following data integration of the PBMC nuclei, unsupervised clustering identified 20 main clusters and 27 subclusters across the dataset (Supplemental Data 3-4). All major expected immune cell types were identified (Figure 2 and S2), including B cells (BACE2, WDFY4, CCSER1), multiple CD4⁺ T cell subtypes (LEF1), CD8⁺ T cells (KLRD1, CD8A, CD8B), and monocyte subsets, comprising CD14⁺ monocytes (FCN1, LYZ) and CD16⁺ monocytes (CLEC7A). Dendritic cells were represented by both cDC2 (FCER1A, CCSER1, FLT3) and pDC (IRF8, CD4, LGMN), and additional populations included NK cells (ADGRG5, KLRD1), MAIT cells, double-negative T cells, plasma cells, and platelets. Each cluster was annotated based on canonical marker gene expression, with all identities reviewed and validated manually. Overall, cell type proportions were highly similar between HSCT survivors and controls, with no major shifts in the relative abundance of the major immune populations (Figure S2). A modest expansion of a CD8⁺ T cell population was detected in HSCT participants compared with controls (Figure S2A). As expected, following allogeneic transplantation, PBMCs from HSCT participants were almost exclusively donor-derived, with the exception of a small residual recipient-derived population in one participant (157 nuclei vs. 16,266 donor-derived). The recipient-derived population clusters separately and appears to consist of non-immune cells, for example expressing MYH2, so likely they reflect a sampling artefact (i.e. tissue-contamination in the blood sample).

**Figure 2.**
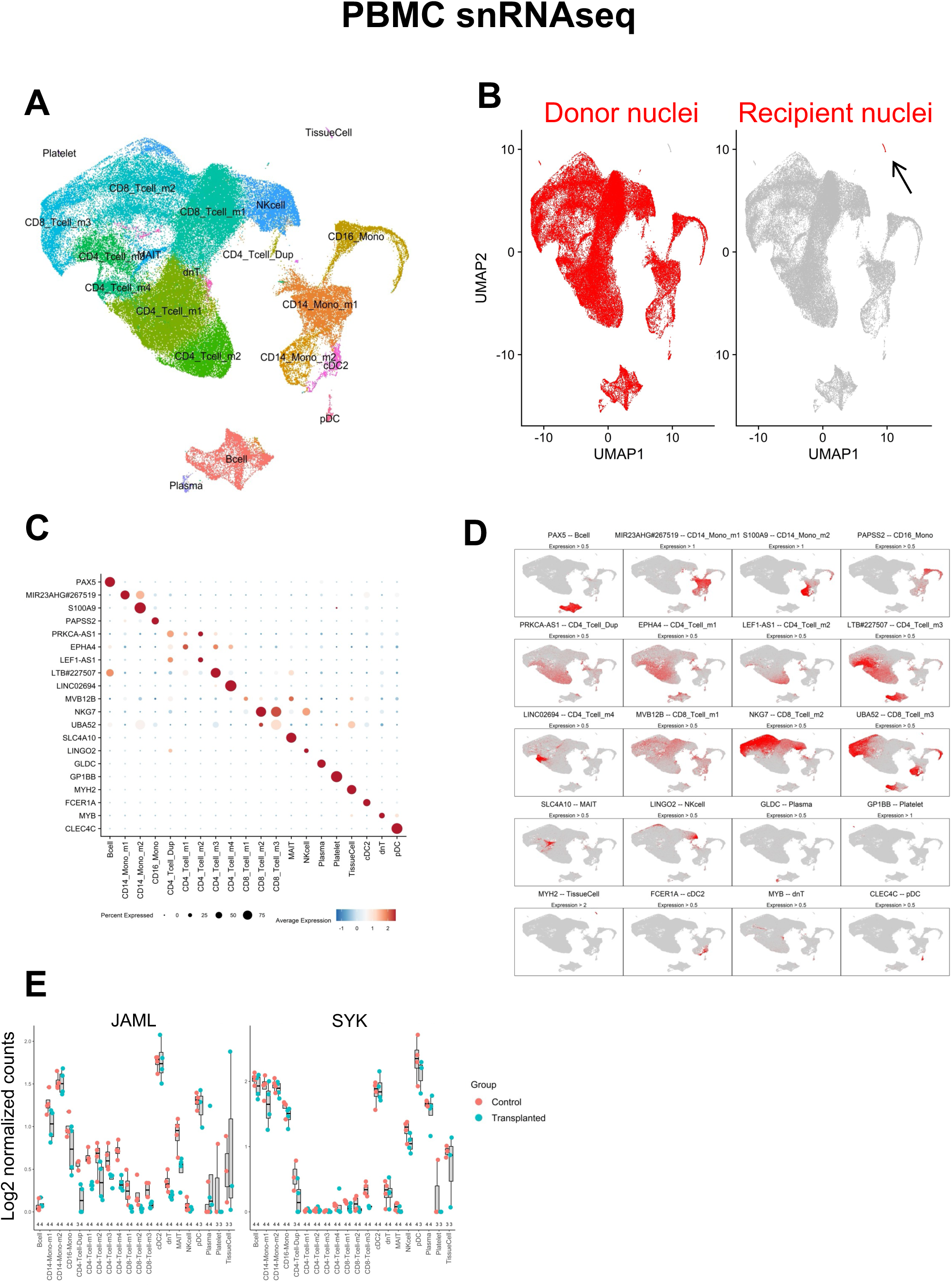
Single-nucleus RNA sequencing of peripheral blood mononuclear cells reveals subtle immune transcriptional alterations in HSCT survivors. Single-nucleus RNA sequencing (snRNAseq) was performed on peripheral blood mononuclear cells (PBMCs) from four HSCT survivors and four matched healthy controls. (A) Uniform manifold approximation and projection (UMAP) embedding of all PBMC nuclei, showing unsupervised clustering and identification of major immune cell populations. (B) UMAP from HSCT participants only, illustrating donor–recipient tracing based on single-nucleotide polymorphisms (SNPs); donor-derived nuclei are shown on the left and recipient-derived nuclei on the right. The arrow indicates a small cluster of tissue-derived cells, likely representing a sampling artefact. (C) Dot plot display of canonical marker genes across the 20 main clusters, confirming accurate cluster annotation. (D) Mini-UMAP feature plots illustrating expression of canonical marker genes used to annotate the 20 main immune cell clusters. (E) Expression of JAML and SYK within each nuclear cluster, the only 2 differentially expressed genes between HSCT survivors and controls, highlighting the overall modest transcriptional differences across immune cell populations.

Differential expression analysis identified four differentially expressed genes (DEGs) in the PBMC dataset (Supplemental Data 5 and 6). At the main cluster level, JAML and SYK were significantly lower in HSCT participants within CD4⁺_Tcell_m1 and CD8⁺_Tcell_m3 clusters, respectively (Figure 2E). At the subcluster level, SSBP2 and APP were similarly downregulated in the CD8⁺_Tcell_s3 cluster. Among these, JAML displayed the most consistent differences across clusters, whereas SYK is of particular clinical relevance as a potential therapeutic target in chronic graft-versus-host disease. JAML encodes a co-stimulatory adhesion molecule expressed in activated T cells and monocytes, where it contributes to immune surveillance and T cell activation^32^. Reduced JAML expression in HSCT may therefore reflect impaired co-stimulatory capacity or dampened peripheral T cell responsiveness. SYK, a non-receptor tyrosine kinase, mediates antigen receptor signaling across multiple immune lineages, including T, B, NK, and myeloid cells, and plays a central role in immune activation and inflammation^33^. Its reduced expression in the CD8⁺_Tcell_m3 cluster may indicate subtle suppression of cytotoxic signaling pathways. Taken together, group differences in PBMC transcriptional profiles were limited, and the overall immune cell landscape remained largely indistinguishable between HSCT survivors and healthy controls.

### No evidence of bone marrow derived cells adopting alternative cell fates in human skeletal muscle tissue

We next analyzed skeletal muscle snRNAseq profiles from vastus lateralis biopsies to determine whether donor-derived hematopoietic cells adopt alternative cell fates in human muscle.

Each participant underwent bilateral muscle biopsies 7 days after a single unilateral resistance exercise bout, yielding a total of 16 muscle samples (8 pairs of rested and exercised samples). Following integration of all nuclei transcriptomes, unsupervised clustering identified 15 main clusters and 63 subclusters across all samples (Figure 3, Figure S3 and Supplemental Data 7 and 8). Myonuclei, which constituted 78.2% of all nuclei, were segregated into 9 clusters (designated Myo_m1-9). Four clusters corresponded to type I myonuclei (MYH7), two clusters to type IIa myonuclei (more MYH2, less MYH1) and one cluster to type IIx myonuclei (more MYH1, less MYH2). Myo_m8 could not be clearly assigned to a specific muscle fiber type, whereas Myo_m9 contained nuclei expressing genes related to neuromuscular junctions and denervation (TAFA4, RYR2, NGS2, MUSK and NCAM1)^34–36^. Noticeably, all myonuclei clusters were present in all participants, and the overall percentage of myonuclei relative to all nuclei is similar to published snRNAseq studies of human vastus lateralis and semitendinosus muscle^37–40^. All remaining nuclei were assigned to distinct mononuclear cell populations commonly found in human skeletal muscle using sn/scRNAseq^34,37–48^. These included adipocytes (Adipo; 0.2%; PLIN4, FASN), endothelial cells (Endo; 8.0%; VWF, SHANK3), fibroblasts (Fib; 5.0%; PDGFRA, DCN), immune cells (Immuno; 0.9%; IKZF1, MS4A4A), muscle stem cells (MuSC; 2.2%; PAX7), and smooth muscle cells (Smooth; 5.4%; EGFLAM). The presence of skeletal muscle transcripts in these mononuclear cell clusters is likely due to muscle fiber cytoplasmic RNA captured in vesicles during the nuclear isolation procedure. The relative proportions of major and subclustered cell types did not differ significantly between HSCT survivors and healthy controls. All participants exhibited the full spectrum of mononucleated cell types, with the exception of one participant who lacked detectable adipocytes. Notably, another participant showed an overrepresentation of adipocytes (>70% of adipocytes in the entire dataset).

**Figure 3.**
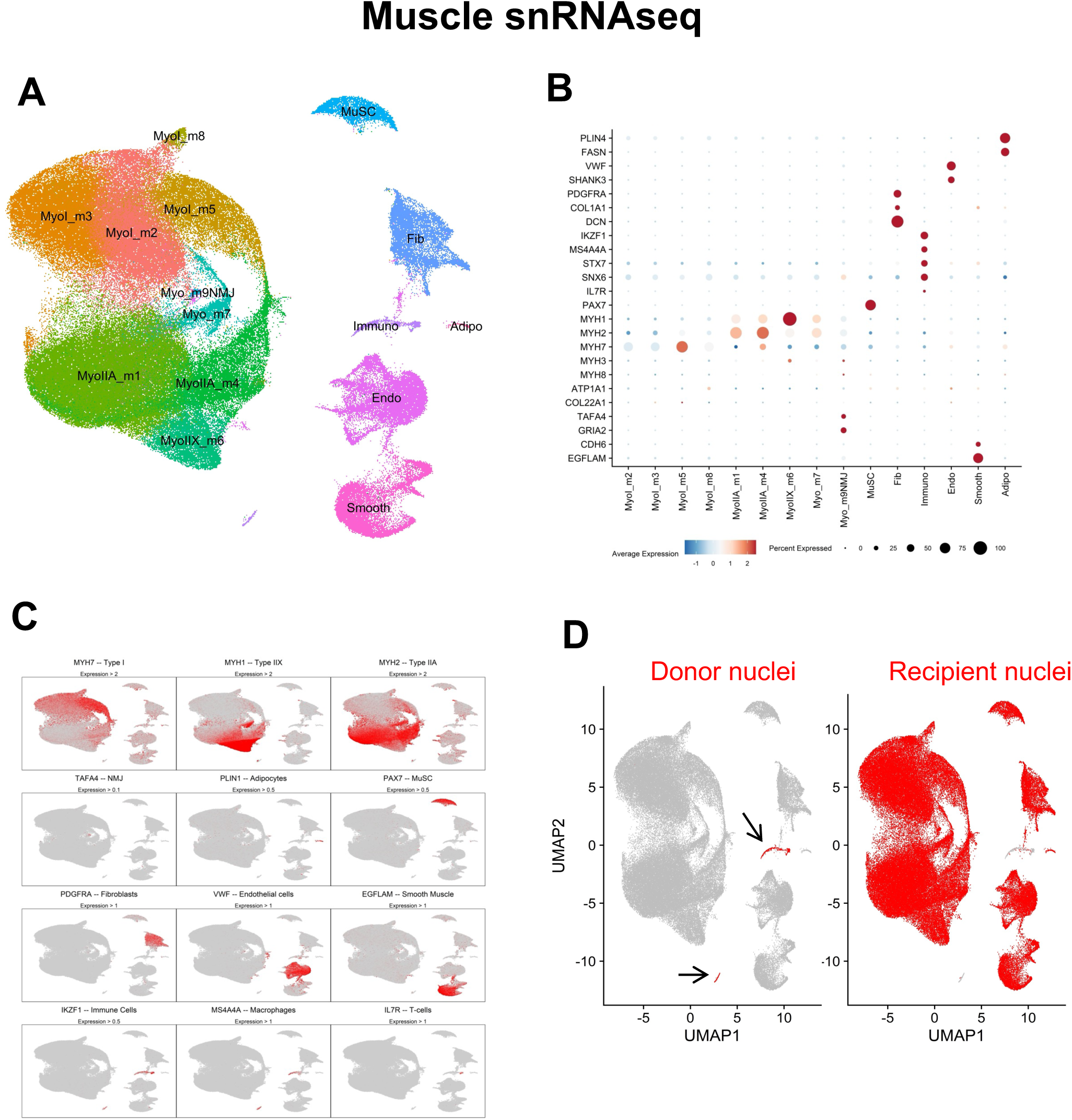
No evidence of bone marrow derived cells adopting alternative cell fates in human skeletal muscle tissue. Single-nucleus RNA sequencing (snRNAseq) was performed on vastus lateralis muscle biopsies obtained from four HSCT survivors and four matched healthy controls. For each participant, biopsies were collected seven days after a single bout of unilateral resistance exercise from the exercised leg and the contralateral leg, yielding 16 samples in total. (A) Uniform manifold approximation and projection (UMAP) embedding of all muscle nuclei, showing unsupervised clustering into 15 major cell populations. (B) Dot plot summarizing canonical marker gene expression across all major clusters, confirming cluster identities. (C) Mini-UMAP feature plots illustrating expression of canonical marker genes used to annotate the major muscle-resident cell populations. (D) UMAP from HSCT participants only, illustrating donor–recipient tracing based on single-nucleotide polymorphisms (SNPs); donor-derived nuclei are shown on the left and recipient-derived nuclei on the right. Only a single donor-derived nucleus was detected within myonuclear clusters, with no donor-derived nuclei identified in other muscle-resident cell populations.

To explore the fate of bone marrow-derived cells in muscle, we used single nucleotide polymorphism (SNP)-based genotyping as the lineage-tracing strategy, which provided robust donor–recipient classification in 99.9 % of nuclei. This approach revealed that all donor-derived nuclei were immune cells (Figure 3D), with the exception of a single donor-derived myonucleus. No donor-derived nuclei were identified in other mononucleated cell populations. In contrast, 8.7% of immune nuclei remained recipient-derived, indicating incomplete turnover of recipient tissue-resident immune cells in human skeletal muscle.

### XRRA1 is upregulated in skeletal muscle and cultured muscle stem cells from HSCT survivors

To assess transcriptional differences in skeletal muscle tissue between HSCT and control participants, we performed differential expression analysis of the snRNAseq muscle dataset across major cell clusters (Figure 4; Supplemental Data 9 and 10). This analysis identified three genes with significant group differences (adjusted p < 0.05): EMC7 (ER membrane protein complex subunit 7, also known as C15orf24), ATP6V0B, and XRRA1 (X-Ray Radiation Resistance Associated 1) (Figure 4). EMC7 was significantly lower in MyoI_m3 of HSCT survivors, with a similar trend across other myonuclear clusters. Of note, EMC7 localizes to the sarcolemma of the neuromuscular junction^49^, and its lower expression in HSCT muscle may suggest subtle disadvantageous neuromuscular remodeling in this cohort. ATP6V0B, which encodes a component of the vacuolar ATPase proton pump, was lower in MyoIIA_m1 of HSCT survivors, again with consistent trends across most myonuclear clusters. In contrast, in the same myonuclei cluster, XRRA1 was significantly upregulated in HSCT survivors with a similar pattern across all other clusters (except adipocytes).

**Figure 4.**
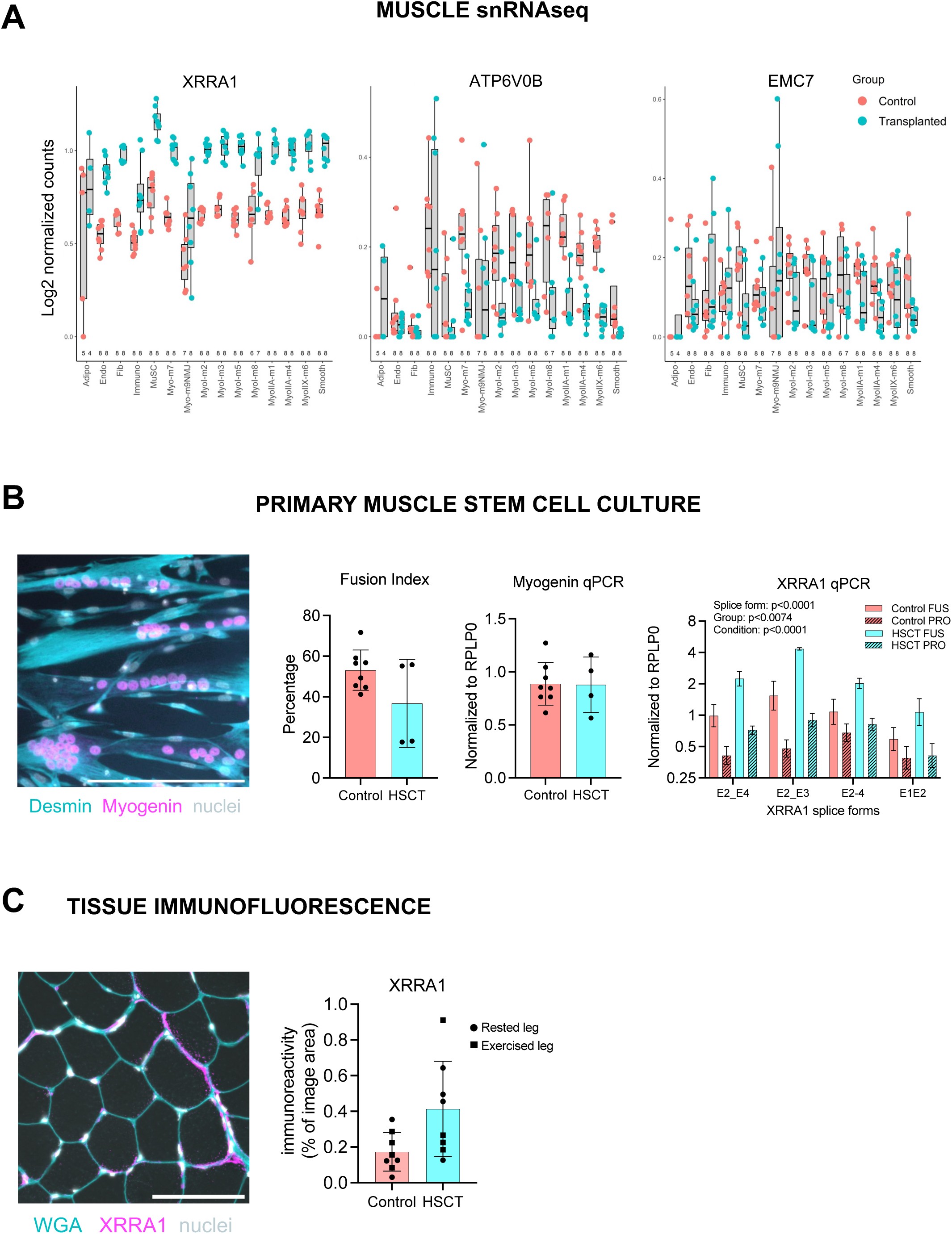
Elevated XRRA1 expression in skeletal muscle tissue and muscle stem cell cultures from HSCT survivors. (A) Differentially expressed genes identified by single-nucleus RNA sequencing (snRNAseq) across myonuclear clusters, highlighting EMC7, ATP6V0B, and XRRA1 as significantly altered in HSCT survivors compared with controls. Data are shown in boxplots. (B) Primary muscle stem cell culture (controls: n = 8; HSCT: n = 4). Representative immunofluorescence image of differentiated myotubes from an HSCT survivor, stained for desmin (cyan), myogenin (magenta) and nuclei (white). Fusion index (percentage of nuclei located within multinucleated (≥3 nuclei) desmin-positive myotubes relative to total desmin-positive nuclei). Quantitative PCR (qPCR) data (geometric mean ± SEM) of myogenin. and XRRA1. XRRA1 data were analyzed across three splice forms, two cellular conditions (proliferating cells (PRO) and fusing cells (FUS)), and group (control vs. HSCT) by three-way repeated-measures ANOVA. (C) Representative immunofluorescence image of a muscle biopsy cross-section from a HSCT survivor stained with wheat germ agglutinin (cyan), XRRA1 (magenta), and nuclei (white). Quantification of XRRA1 fluorescence intensity is shown for rested (circles) and exercised (squares) legs from control (4 rested / 4 exercised) and HSCT participants (4 rested / 4 exercised); mean ± SD with individual values.

XRRA1 is implicated in the cellular response to DNA damage, although its precise function remains poorly characterized. Initially identified in 2003^50^, XRRA1 was named based on findings that X-ray exposure increased its expression in radiosensitive tumor cells, suggesting a potential role in DNA damage response pathways^51^. While this aligns with its putative function in cancer contexts, more recent studies indicate a more complex picture. In a single-cell RNAseq study of mouse skeletal muscle, XRRA1 expression was not induced at either 24 hours or 56 days following X-ray exposure^52^. Conversely, XRRA1 has been reported to be markedly upregulated in blood samples of leukemia patients prior to any treatment^53^, suggesting links to underlying hematologic malignancy and/or therapy-induced stress responses, and likely explaining the persistent XRRA1 upregulation observed in skeletal muscle years after allogeneic HSCT. The strong and consistent XRRA1 upregulation in our data prompted further exploration on tissue sections and in primary muscle stem cell cultures.

To investigate XRRA1 expression at the protein level, we performed immunofluorescence staining on skeletal muscle cross-sections (Figure 4C and S4). XRRA1 signal was detectable in all samples and localized predominantly within muscle fibers, with the strongest staining observed in discrete sarcolemmal patches. Quantification of XRRA1-positive pixel area indicated approximately two-fold higher XRRA1 signal in the muscle cross-sections of HSCT survivors compared with controls (Figure 4C).

To explore whether XRRA1 expression would remain elevated in cells upon removal from their native environment, as demonstrated in other contexts^54^, we isolated primary muscle stem cells from the vastus lateralis biopsy tissue from 4 HSCT participants and 8 healthy controls. Muscle stem cells were cultured under proliferation and fusion conditions (Figure 4B). Three XRRA1 splice forms and myogenin expression levels were quantified by qPCR (Figure 4B). A three-way repeated-measures ANOVA with factors Group (HSCT, control), Condition (proliferation, fusion), and XRRA1 Splice form revealed a main effect of Group (p < 0.01), indicating higher XRRA1 expression in MuSCs from HSCT survivors across splice forms and stage of myogenesis. Additional main effects of Condition (p < 0.0001) and Splice form (p < 0.0001) were observed. Fusion index and myogenin gene expression were similar between groups, confirming that the differences in XRRA1 were not attributable to compromised myogenic potential. Taken together, these findings, in line with the snRNAseq data, indicate that elevated XRRA1 expression represents a persistent, cell-intrinsic imprint on MuSCs from HSCT survivors, even after multiple rounds of cell division outside the native environment.

Consistent with the tissue-level upregulation of XRRA1, the PBMC snRNAseq data revealed a modest trend toward higher XRRA1 expression across most immune cell clusters, although no single cluster reached statistical significance (Figure S2). This suggests that XRRA1 dysregulation is also present in donor cells transplanted after radiation and chemotherapy, indicating a lasting effect of treatment on the cell niche.

At the subcluster level, 41 genes were differentially expressed between HSCT survivors and controls in muscle (Supplemental Data 10). Most changes were observed in endothelial, smooth muscle, and myonuclear subclusters, with pathways related to mitochondrial energy metabolism, cellular stress, and immune signaling. Notably, PKHD1L1 and MMRN1 were elevated in the EndoSmooth subcluster, whereas SLC25A5 was reduced in the MyoCyto_s1 cluster. MMRN1, PKHD1L1, and SLC25A5 have all been implicated in chemotherapy resistance and survival pathways in multiple malignancies^55–57^. In smooth muscle cell subclusters, several genes involved in mitochondrial and stress-response pathways (MT-CO3, MT-ND3, CKM, HSP90AA1) were significantly lower in the HSCT group.

### Marked impairments in muscle fiber innervation in HSCT survivors

The observation of downregulated EMC7 expression in HSCT muscle, together with previous reports of neuromuscular complications in allogeneic HSCT recipients^8^, prompted us to examine potential alterations in muscle fiber innervation status. We assessed four complementary histological and systemic markers of denervation and neuromuscular junction integrity (Figure 5A-D). Firstly, HSCT muscle displayed a significantly higher prevalence of fiber type grouping (p < 0.05)^58^, consistent with repeated cycles of denervation and reinnervation, or failed reinnervation leading to eventual loss of denervated fibers. Secondly, the frequency of central nuclei, which are associated with denervation in various neurogenic disorders^59^, was significantly higher in HSCT muscle (p < 0.01). Thirdly, NCAM immunofluorescence was used to identify denervated muscle fibers^35^, where a trend toward a higher proportion of NCAM⁺ fibers in HSCT muscle compared with controls (p = 0.0549) was observed. In addition, circulating C-terminal Agrin Fragment (CAF), a biomarker of neuromuscular junction instability^60^, was significantly elevated in HSCT survivors (p < 0.001, Figure 5B). Finally, to integrate these observations, Z-scores were calculated for each marker and averaged to derive a composite Denervation Score, which was significantly higher in HSCT survivors than in controls (p < 0.0001, Figure 5C), providing converging evidence of impaired neuromuscular integrity in HSCT survivors, characterized by increased markers of denervation, altered muscle fiber organization, and elevated circulating indicators of neuromuscular junction instability.

**Figure 5.**
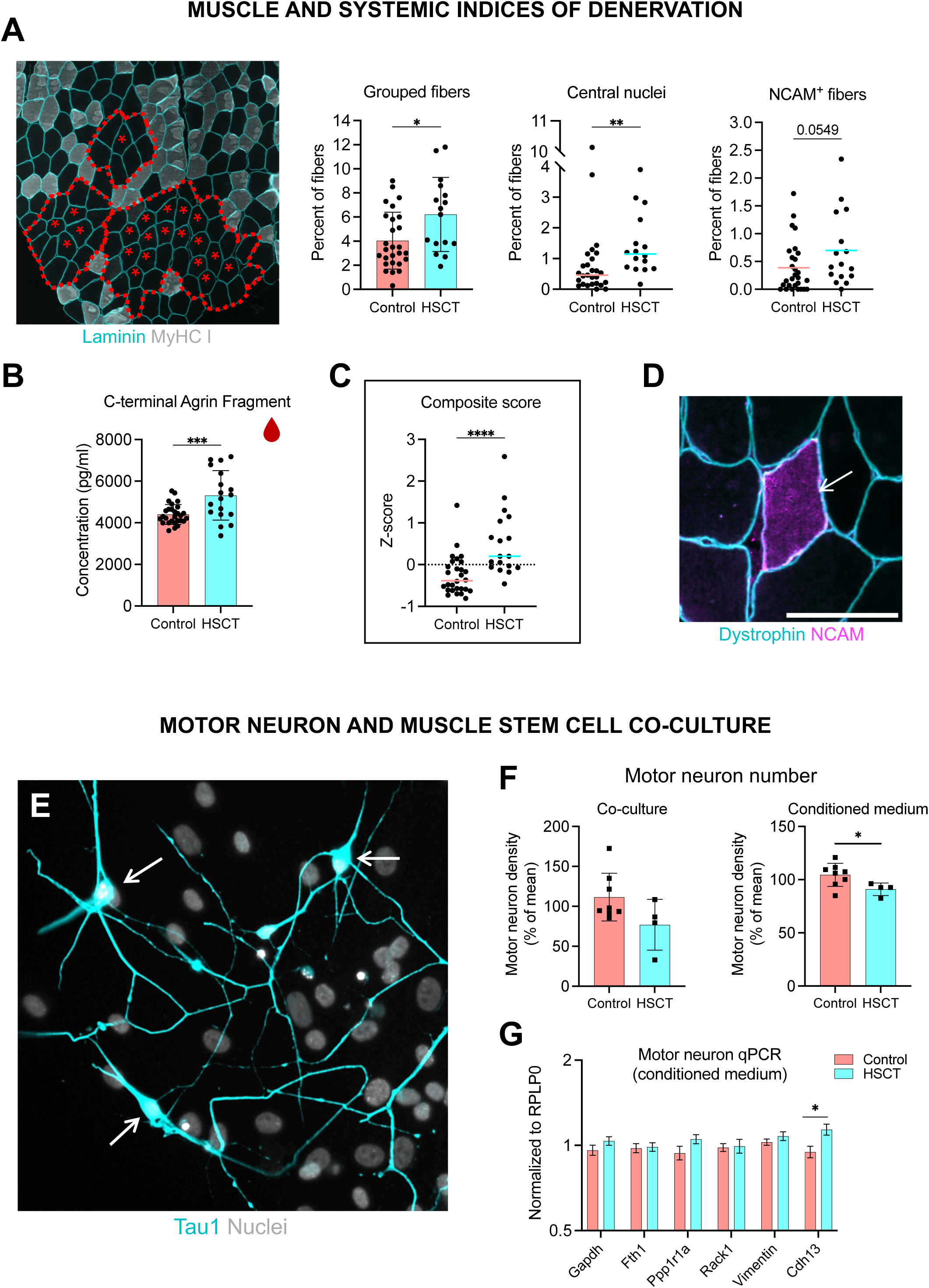
Marked impairments in muscle fiber innervation and motor neuron support in HSCT survivors. (A) Muscle markers of denervation. Grouped fibers (controls: n = 27; HSCT: n = 16; unpaired two-tailed t-test; mean ± SD) as seen in representative immunofluorescence image, centrally nucleated fibers (controls: n = 27; HSCT: n = 16; Mann–Whitney U-test, median value indicated), and NCAM-positive fibers (controls: n = 28; HSCT: n = 16; Mann–Whitney U-test, median value indicated). (B) Serum levels of the C-terminal agrin fragment (CAF) (controls: n = 28; HSCT: n = 18; unpaired two-tailed t-test; mean ± SD). (C) Composite denervation score expressed as Z-scores averaged across all innervation indices (A–B) (controls: n = 28; HSCT: n = 18; Mann–Whitney U-test, median value indicated). (D) Representative immunofluorescence image of an NCAM-positive fiber (scale bar = 100µm), stained for dystrophin (cyan) and NCAM (magenta), for quantification of data in A. E-G Motor neuron and muscle stem cell co-culture (E) Representative image of rat primary motor neurons cultured on human muscle cells (scale bar = 100µm), stained for Tau1 (cyan) and nuclei (white). (F) Quantification of motor neuron survival in functional co-culture assays, either direct cell–cell contact or indirect exposure via conditioned medium (controls: n = 8; HSCT: n = 4; mean ± SD). (G) qPCR of selected neurite-associated genes in motor neurons following exposure to conditioned medium derived from control and HSCT muscle cells (controls: n = 8; HSCT: n = 4; geometric mean ± SEM). Statistical significance is indicated as *p < 0.05, **p < 0.01, ***p < 0.001, and ****p < 0.0001.

To further assess the functional impact of HSCT on neurotrophic action, we performed co-culture experiments using primary human MuSCs, and motor neurons isolated from E15 Sprague Dawley rat embryos^61^. Both direct co-culture and indirect exposure via conditioned medium were employed to evaluate motor neuron viability, quantified by ChAT and Tau1 immunofluorescence (Figure 5E). Motor neuron survival in direct co-culture with HSCT-derived MuSCs was not inferior to that observed with control MuSCs (Figure 5F). In contrast, exposure to conditioned medium from HSCT-derived MuSCs led to a significant reduction in motor neuron viability within 48 hours (p < 0.05; Figure 5F). At the gene-expression level, Cdh13, a negative regulator of neurite growth and axon guidance^62^, was higher in motor neurons exposed to conditioned medium from HSCT-derived MuSCs (p < 0.05; Figure 5G), indicating a delay in neurite growth^63^. These findings suggest that secreted factors released by HSCT MuSCs negatively influence motor neuron activity and survival, pointing to a cell-extrinsic mechanism that may contribute to the neuromuscular innervation deficits observed in HSCT survivors.

### Subtle transcriptional differences in response to a single exercise bout in HSCT survivors

We next examined the acute muscle response to a single bout of unilateral HReT (Figure 6, Figure S5, and Table S1). The exercise protocol consisted of maximal concentric and eccentric contractions performed at both slow (30°/s) and fast (180°/s) angular velocities. HSCT survivors and healthy controls exerted similar relative forces and reported comparable ratings of perceived exertion (RPE), indicating that participants in both groups were able to perform the high-intensity exercise bout to a similar degree. As expected, maximal voluntary contraction (MVC) was acutely reduced in both groups following exercise (-21 ± 9 %; data not shown), consistent with the development of fatigue, and no significant between-group differences were observed.

**Figure 6.**
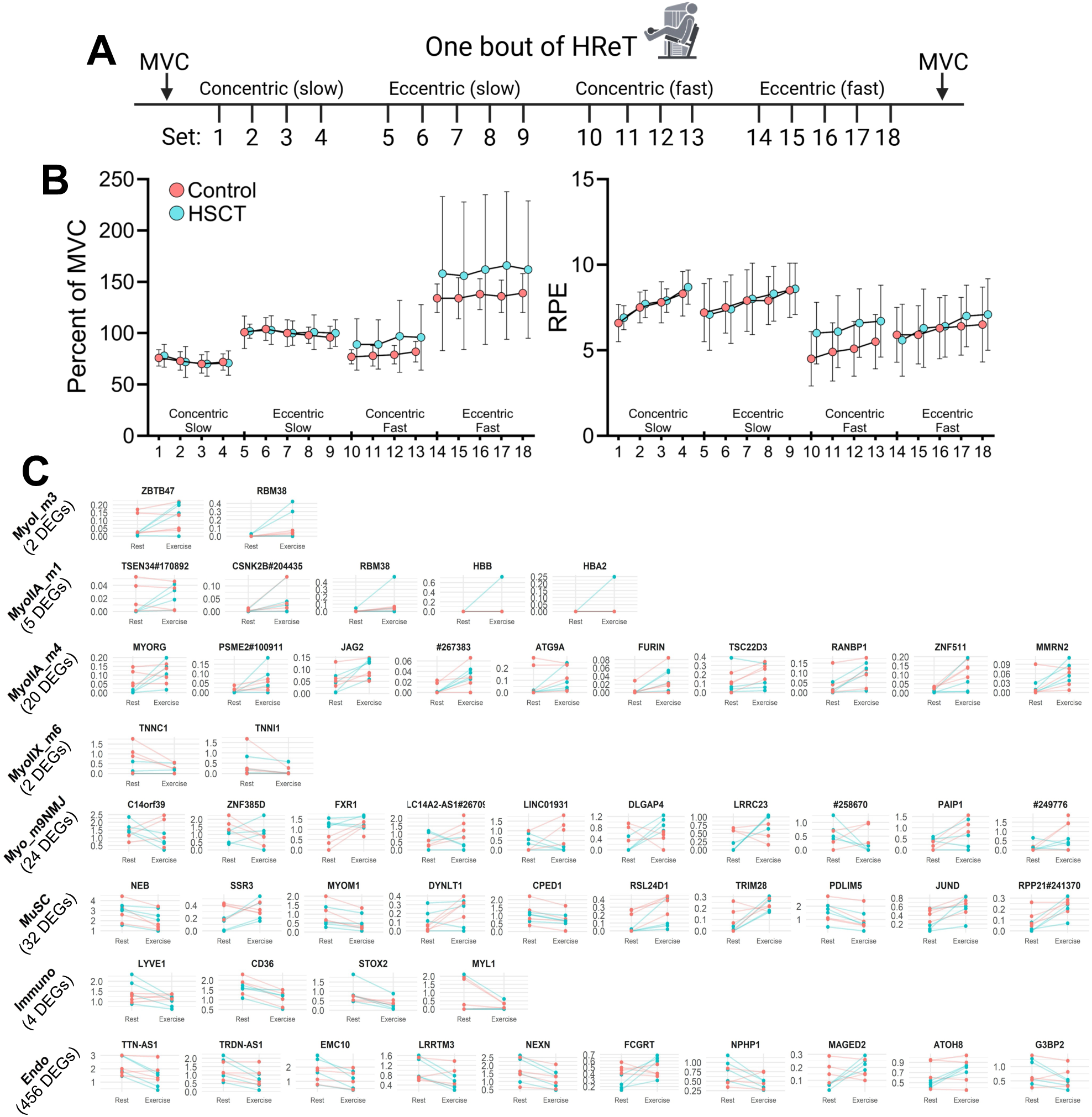
Subtle transcriptional differences in the skeletal muscle response to a single bout of heavy resistance exercise in HSCT survivors. (A) Schematic overview of the single bout unilateral resistance exercise protocol, consisting of maximal concentric and eccentric contractions performed at slow (30°/s) and fast (180°/s) angular velocities. Muscle biopsies were obtained from both the rested and exercised legs seven days after the exercise bout. (B) Relative force output during the single exercise bout, expressed as a percentage of maximal voluntary contraction (MVC) for each contraction velocity. Force output represents the average of the first, middle, and last repetition of each set. Ratings of perceived exertion (RPE) were recorded after each set (controls: n = 11; HSCT: n = 7). Data are mean ± SD. C) Cell clusters exhibiting differential transcriptional responses to exercise between groups, based on significant group × exercise interaction effects. The top 10 differentially expressed genes (DEGs) ranked by log₂fold change are shown for each cluster, with the total number of DEGs indicated below. See Supplemental Figure 5 for the complete set of exercise-responsive DEGs (main effect of exercise).

Muscle biopsies were collected simultaneously from the rested and exercised legs, seven days after the single exercise bout. This design allowed for a within-subject comparison of post-exercise tissue responses while minimizing potential circadian or temporal confounders, as reported elsewhere^64^. The snRNAseq main cluster data revealed a higher proportion of immune cells across both groups in the exercised leg (Figure S3), along with a trend toward increased smooth muscle cell abundance. At the subcluster level, the proportions of five subpopulations, primarily endothelial, smooth muscle, and fibroblast lineages, were increased in the exercised leg (not shown).

To assess transcriptional responses to exercise, we compared exercised vs. rested legs across 15 main cell clusters and identified 1120 DEGs (Figure S5; Supplemental Data 11 and 12). The top 5 largest responses were observed in the Endo, MyoIIA_m4, MuSC, MyoI_m5 and Fib clusters. Endo showed a particularly strong response (877 DEGs; 478 down and 399 up), with GO–Biological Process enrichment for circulatory and cardiac pathways (Supplemental Data 13-16), and modest downregulation of XRRA1. MyoIIA_m4 (77 DEGs; 1 down and 76 up) and MyoI_m5 (29 DEGs; 1 down and 28 up) exhibited transcriptional signatures (represented by e.g. SYN3, LAMA5, TIAM1, DCC, ROR2, AP2M1, SCRIB, PTPRT2, CDH12, DPP10, CTNND2, DSCAM, SHANK3, CNTNAP2) related to synapse organization, cell adhesion, and neuronal processes, consistent with exercise-induced strengthening of neuromuscular innervation. MuSCs displayed balanced up- and downregulation (49 DEGs; 30 down and 19 up), with pathways linked to muscle differentiation and structural remodeling, while Fib (23 DEGs; 2 down and 21 up) showed a smaller response without a dominant functional theme. These findings align with previous sc/snRNAseq studies of acute exercise in human skeletal muscle^42,43,65,66^, while extending the follow-up to 7 days, later than in earlier reports (3–24 h), and providing the first circadian-controlled muscle snRNAseq dataset. In a recent study using bulk muscle tissue RNAseq, a large proportion of the 2399 DEGs in the 24-h post exercise window related to transcription, translation, and the synthesis of new ribosomes, as well as demonstrating a wide range in log2 fold changes from baseline^64^. While our snRNAseq data represent pre-RNA, in contrast to the mRNA measured by bulk tissue RNAseq, we find it noteworthy that such a broad response across cell clusters is still evident 7 days after exercise, and the small fold changes with little overlap in GO terms (relative to the study by Edman and colleagues) at this time point is in line with the dynamic temporal pattern of adaptive processes involved in muscle adaptation.

As our primary focus, we assessed Group × Exercise interaction effects to determine whether the transcriptional response to acute exercise differed between HSCT survivors and controls (Figure 6; Supplemental Data 17 and 18). This analysis identified 545 DEGs, with the largest proportion of DEGs observed in Endo (456 DEGs), followed by MuSCs (32 DEGs), Myo_m9NMJ (24 DEGs), and MyoIIA_m4 (20 DEGs). Most interaction DEGs displayed modest log2-fold differences, indicating subtle transcriptional differences between groups. Nevertheless, several patterns were noteworthy. Selected genes involved in mechanosensing and angiogenic signaling (NPHP1, NEK10, ESR1)^67–70^, as well as immune or stress-related pathways (FCGRT, ATOH8, MAGED2, SERPINH1)^71–73^, exhibited differential regulation between HSCT survivors and controls. In MuSCs and Myo_m9NMJ, differences were small, whereas substantial differences were observed in TSEN34 (MyoIIA_m1), MYORG (MyoIIA_m4), TNNC1 (MyoIIX_m6), and ZBTB47 (MyoI-m3), a zinc-finger transcription factor implicated in neurodevelopmental signaling^74^. Interestingly, the some of these genes stand out with their large fold differences between groups. For example, the upregulation of TSEN34 in HSCT survivors with exercise was ∼48x (5.6 log2FC) that of controls, while the difference for MYORG and ZBTB47 was ∼3x (1.7 log2FC) and ∼5x (2.5 log2FC), respectively. However, it should be noted that these seemingly large differences in upregulation between the groups were often driven by lower levels in the HSCT control leg (Figure 6).

Given the positive response of HSCT survivors to a single bout of exercise, we proceeded with a 12-week training intervention aimed at restoring the impaired muscle mass and function in the HSCT survivors.

### Preserved capacity for responding to heavy resistance exercise training

To evaluate the adaptive capacity of HSCT survivors to heavy resistance exercise training (HReT), seven patients and sixteen healthy controls completed a 12-week supervised training program (3 sessions per week) (Figure 7, table S4). *In vivo* physiological testing and muscle biopsies were performed before and after the intervention. The training protocol has been described in detail previously^75^, and the primary outcome of the trial (ClinicalTrials.gov: NCT04922970), reported here for the first time, was the change in quadriceps cross-sectional area (CSA) assessed by MRI.

**Figure 7.**
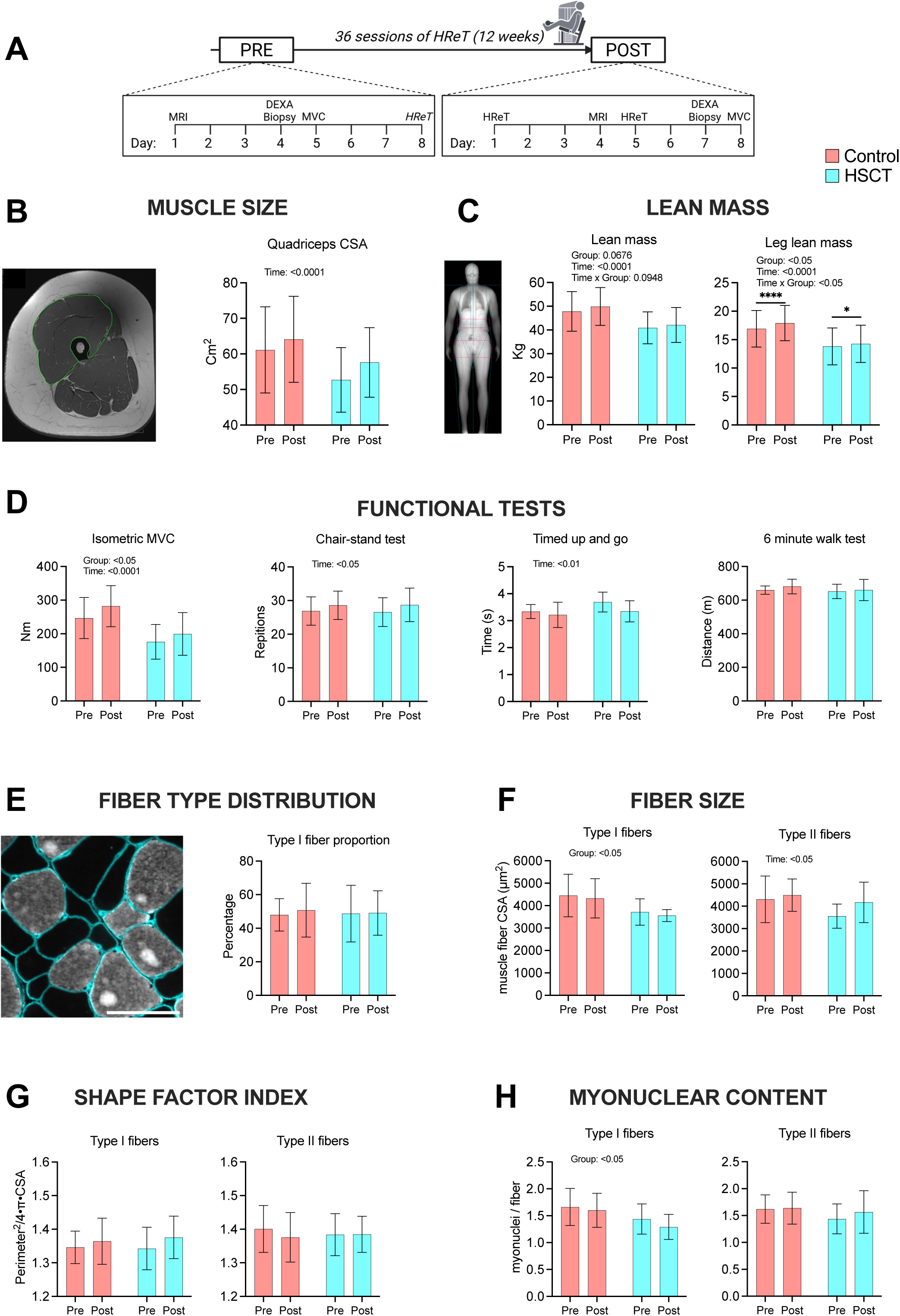
Preserved physiological adaptation to 12 weeks of heavy resistance exercise training in HSCT survivors. Physiological and cellular adaptations to a 12-week heavy resistance exercise training (HReT) program were assessed using two-way repeated-measures ANOVA with factors Group and Time. Data are mean ± SD. (A) Schematic overview of the 12-week supervised HReT intervention and timeline of pre- and post-intervention testing and muscle biopsies. (B) Representative magnetic resonance imaging (MRI) of thigh used to quantify quadriceps muscle cross-sectional area (CSA) (controls: n = 16; HSCT: n = 6). (C) Whole-body and leg lean mass assessed by DXA (controls: n = 15; HSCT: n = 6) (D) Isometric maximal voluntary contraction (MVC) assessed by dynamometry (controls: n = 15; HSCT: n = 7) and functional performance assessed using the 30-s chair-stand test, timed up-and-go test, and 6-min walk test (controls: n = 10; HSCT: n = 7). (E) Fiber type I proportion (controls: n = 13; HSCT: n = 7) (F) Cross-sectional area (CSA) of type I and type II fibers (controls: n = 13; HSCT: n = 7). (G) Shape factor index of type I and type II fibers (controls: n = 13; HSCT: n = 7). (H) Myonuclear content of type and II fibers (controls: n = 13; HSCT: n = 7).

Both groups demonstrated substantial increases in quadriceps muscle size over time (p < 0.0001), with no Group × Time interaction, indicating similar training-induced hypertrophy in HSCT survivors (9.3 %) and controls (4.9 %; Figure 7B). Whole-body and leg lean mass increased over time (both p < 0.0001; Figure 7C). Although a Group × Time interaction was detected for leg lean mass, the absolute difference in trajectory was small and did not alter the overall conclusion that both groups responded robustly to training (5.9 % in controls, 3.2 % in HSCT). Isometric maximal voluntary contraction (MVC) improved significantly following training (p < 0.0001), again with comparable strength gains in both groups (14.3 % in controls, 13.3 % in HSCT; Figure 7D). Muscle biopsy analyses revealed significant hypertrophy of type II fibers (p < 0.05; 4.4 % in controls, 17.3 % in HSCT), with no changes in type I fibers (Figure 7F). Fiber type proportions (Figure 7E), fiber type-specific SFI (Figure 7G) and myonuclear content (Figure 7H) remained unchanged in both groups. Main effects of time were observed for one-repetition maximum (1RM) in leg extension, leg press, and leg curl, with no Group × Time interactions (Figure S6). Health-related quality of life (SF-36) and resting blood pressure parameters did not change in either group over the intervention period and remained within normal ranges (data not shown). Taken together, these findings demonstrate a preserved ability of HSCT survivors to respond to resistance training, achieving muscle mass and strength gains comparable to those of healthy controls.

## DISCUSSION

Our findings reveal that long-term survivors of pediatric HSCT carry lasting molecular imprints of cancer-disease and its treatment on their skeletal muscles, in line with the lower muscle mass and function compared to healthy counterparts. However, most importantly they retain a preserved capacity to respond and adapt to both a single bout of heavy resistance exercise training (HReT) and to 12 weeks of regular HReT, overcoming neuromuscular deficits.

HSCT survivors exhibited markedly reduced muscle mass and strength, reflecting profound physiological impairment. At the molecular level, snRNAseq revealed several differentially expressed genes in cell subpopulations of both muscle and PBMCs, with *XRRA1*, a gene linked to DNA damage^50^, being most prominently and broadly upregulated in survivors. In addition, the altered profile for SLC25A5, and MMRN1 and PKHD1L1 in endothelial cells, supports a general picture of a lasting transcriptional imprint of HSCT regimen within skeletal muscle and its vasculature >10 years after treatment. The lower EMC7 expression in myonuclei clusters, together with the elevated negative axon guidance cue Cdh13^62^ in motor neurons exposed to HSCT survivor myotube conditioned medium, are consistent with our histological findings of muscle fiber denervation. Muscle innervation is maintained by both sides of the NMJ, where the motor neuron presynaptic terminal meets the postsynaptic motor endplate, and emerging evidence indicates that exercise has the potential, not only to remodel the NMJ but also to enhance motor neuron survival. For example, exercise attenuates age-related changes in the mouse NMJ^76^, and our previous data in humans indicate that lifelong exercise enhances the neurotrophic potential of muscle fibroblasts and myoblasts^61^. Our conditioned medium findings strongly implicate the muscle compartment as the source of treatment-induced damage driving the impaired innervation, although we cannot rule out additional central effects on the motor neurons. Regardless of the source, once denervated, or when neuromuscular transmission is compromised, muscle fibers rapidly atrophy and undergo disassembly of their contractile machinery and eventually die^77^. Therefore, it is likely that the pronounced weakness in the HSCT survivors is due to a combination of muscle fiber loss and reduced muscle fiber size.

Notably, sex-specific differences were apparent. Although the cohort size precludes definitive conclusions, male survivors appeared more severely affected at the molecular level than their female counterparts. Myonuclear domain, the cytoplasmic territory governed by each nucleus^78^, was lower in male HSCT survivor type I myofibers only compared with controls. Since smaller fibers have smaller domains^79,80^, this result could be explained by the trend for smaller type I myofibers in the male HSCT survivors vs controls. However the smaller domain size, and similar number of myonuclei per fiber, rules out deficiencies in myonuclear accretion as a limiting factor for myofiber growth during the childhood/adolescent growth period after HSCT treatment, in contrast with the mouse prepubertal radiation model demonstrating decreased myofiber size and myonuclear content of both type I and II fibers 14 months post radiation^81^. It is therefore more likely that the lower muscle and fiber size (along with functional outcomes) in our participants is due to low levels of physical activity, rather than defects in physiological adaptation to exercise, as confirmed in our RCT. Nonetheless the loss of muscle fibers through denervation, discussed above, is irreversible and places HSCT survivors on a poorer trajectory, which could reduce healthspan and years of independent living in the last decades of life.

HSCT survivors mounted a robust response to a single bout of exercise, with histological, *in vivo*, and transcriptional adaptations that were broadly comparable to healthy controls. Although some DEGs emerged from the snRNAseq dataset distinguishing the groups, the effect sizes were generally small, indicating that pre-existing treatment imprints did not blunt the acute exercise response. Notably, only four prior studies have applied these technologies to human skeletal muscle in the context of exercise^42,43,65,66^, and this is the first to employ a within-individual design, sampling both the rested and exercised leg simultaneously to minimize circadian and environmental influences. Consistent with earlier reports, the single exercise bout elicited marked transcriptional remodeling across multiple cell types, including endothelial cells, macrophages, fibroblasts, muscle stem cells, and myonuclei, underscoring the broad cellular participation in the adaptive response. We next asked whether the preserved acute responsiveness would translate into long-term plasticity when survivors were challenged with a structured 12-week program of HReT. Despite starting from a lower baseline, HSCT survivors showed relative improvements in multiple indices of muscle mass and strength comparable to controls, including the primary outcome of quadriceps size assessed by gold-standard MRI. This finding has profound clinical implications. First, it directly contrasts with the only prior study of long-term survivors undergoing HReT^14^, which, lacking a control group, reported no gain in lean body mass after six months, suggesting that adaptability was impaired. Second, even though exercise can be implemented in close proximity to transplantation^10–13^, our data demonstrate that rehabilitation at a later stage still permits recovery of muscle mass and strength. However, whether the greater rate of muscle fiber denervation in HSCT survivors prevents them from fully recovering lost muscle mass remains to be shown. Since there is no evidence for replacement of lost muscle fibers in adult human skeletal muscle, the fibers lost through denervation likely leave this population at a permanent disadvantage as they age. Initiating exercise training earlier rather than later may therefore attenuate denervation and thereby preserve muscle fiber number.

Addressing a long-standing question in muscle biology, we find no evidence that bone marrow–derived immune cells adopt alternative fates in humans. Using SNP-based donor–recipient tracing, we detected only a single donor-derived myonucleus across 79,315 myonuclei in HSCT-survivors, while donor origin was otherwise restricted to immune clusters. In mice, bone marrow–derived cells have been shown to enter the myogenic lineage, fuse with myofibers during regeneration or disease, restore dystrophin in mdx muscle after transplantation, and even transiently adopt a satellite cell–like state following injury^25,82–84^. In humans, Y-chromosome fluorescence *in situ* hybridization of skeletal muscle tissue 6–12 years post-transplant from sex-mismatched HSCT survivors revealed a Y⁺ nucleus in 0.5–1.0% of myofibers and endothelial cells, interpreted as evidence of rare fusion-mediated donor incorporation^29^. Extrapolating this frequency to our dataset, ∼460–920 donor-derived nuclei would be expected among the 92,459 myonuclear and endothelial nuclei we sequenced. Instead, we identified only a single donor-derived myonucleus, suggesting that such events are not a prominent physiological feature in human skeletal muscle that has undergone childhood/pubertal growth and during homeostatic conditions spanning several decades. Instead, the persistence of recipient-derived macrophages highlights the long-lived nature of resident immune cell populations in human skeletal muscle and underscores important species differences in how bone marrow–derived cells contribute to muscle growth and maintenance.

Beyond these biological insights, this study provides a powerful resource for pediatric hematology and oncology as well as cancer survivorship research. By combining skeletal muscle and peripheral PBMCs, we generated an snRNAseq dataset that enables exploration of HSCT-associated gene expression differences as well as the muscle response to HReT at 7 days when the initial stress response has subsided and more long term adaptation is ongoing. Importantly, we also provide what is, to our knowledge, the first snRNAseq dataset of PBMCs^85^. While scRNAseq is traditionally used for blood profiling, snRNAseq offers complementary advantages, particularly when nuclear transcripts are of interest, and ensures comparable profiles to tissues processed in the same manner. Together, these datasets establish a benchmark resource for mechanistic discovery in pediatric hematology and oncology, skeletal muscle biology, and systemic immune regulation, with broad relevance for understanding the long-term sequelae of cancer treatment and the impact of exercise in mitigating these effects.

This study has some limitations. The cohort size was modest, with an uneven distribution of sexes, which restricts the power to detect sex-specific differences. Ideally, all participants would have contributed to the baseline, single bout, and 12-week intervention arms; however, the rarity of this clinical population and the need to minimize participant burden limited the feasibility of such a design. Despite these constraints, the integration of physiological, histological, and transcriptomic data across multiple experimental contexts provides a uniquely comprehensive and translational view of skeletal muscle adaptation after HSCT.

In summary, our findings reveal that long-term survivors of pediatric HSCT carry persistent molecular and structural imprints of cancer and its treatment, yet they retain a remarkable capacity to regain muscle mass and strength when challenged with heavy resistance training. This delivers a hopeful message for cancer survivorship, that despite enduring treatment-related neuromuscular damage, the physiological response to HReT remains intact, allowing this population to partly recover lost muscle mass and function due to treatment and build up an important reserve for old age.

## MATERIALS AND METHODS

### Study design and setting

This single-center, non-randomized clinical trial investigated the mechanisms underlying impaired muscle function in hematopoietic stem cell transplant (HSCT) survivors and their response to both a single bout and 12 weeks of heavy resistance exercise training (HReT) in two sub-studies. Both studies were approved by the Committee on Health Research Ethics of the Capital Region of Denmark (H-19055648 and H-20076662) and were conducted in accordance with the Declaration of Helsinki. The 12-week training trial was pre-registered at ClinicalTrials.gov (NCT04922970; Figure S7), with change from pre to post in quadriceps cross-sectional area registered as the primary outcome. The study is reported in accordance with the STROBE guidelines^86^.

### Participants

Survivors of pediatric HSCT were recruited through the outpatient clinic at the National Center of Pediatric HSCT at Copenhagen University Hospital, Denmark. All patients had received HSCT before the age of 18 years, between 1992 and 2011. In both the single exercise bout trial and the 12-week training trial, adult HSCT female survivors, who were currently living in Denmark, were invited to participate. Exclusion criteria were any illness or physical disability that could hinder participation in the exercise intervention or clinical examinations; pregnancy; chronic anemia; adherence to a strictly vegan diet; or recent involvement in regular, structured physical exercise. A control group matched for sex and age was included in each sub-study.

### Interventions

In the single exercise bout trial, participants attended two visits at the study site. During Visit 1, a blood sample was collected, followed by a DXA scan and a maximal voluntary contraction (MVC) strength test. Participants then completed a single bout of unilateral heavy resistance exercise training (HReT), with the exercised leg randomized based on leg dominance; the contralateral leg served as the control. Seven days later, participants returned for Visit 2, which included another blood sample and muscle biopsies from both legs. In the 12-week training trial, participants were assessed before and after a 12-week HReT intervention (Table S4). Pre- and post-intervention testing included a blood sample, a DXA scan, an MRI scan of the quadriceps muscles, a muscle biopsy from the left leg and an MVC test for the right leg. Post-tests were conducted 2–3 days after the final training session. In both sub-studies, participants were instructed to refrain from physical activity for 48 hours prior to testing. Additionally, for the single bout exercise trial, participants were asked to avoid structured exercise between Visit 1 and Visit 2. In the 12-week training trial only, health-related quality of life was assessed using the Short Form-36 (SF-36) questionnaire, and resting blood pressure was measured.

### Assessment of muscle mass and strength

Magnetic resonance imaging (MRI) of the thigh musculature was conducted at the Department of Diagnostic Radiology, Copenhagen University Hospital – Rigshospitalet, using either a Siemens 3.0 T scanner or a General Electric 1.5 T scanner. The same scanner was used for both scans of each participant. A T1-weighted sequence was acquired, and images were obtained using a Dixon protocol. Transversal scans with a slice thickness of 4 mm were acquired every 25 mm along the entire thigh. The slice closest to 50% of femur length was selected for analysis. Image analysis was performed as previously described^87^. Briefly, using the Horos software (Horosproject.org, Nimble Co LLC d/b/a Purview, Annapolis, MD, USA), the muscle boundaries of vastus lateralis, rectus femoris and vastus medialis together with vastus intermedius were manually delineated. Quadriceps CSA was calculated as the sum of these four muscles. All scans for each participant were analyzed together, with the analyst blinded to both time point and group allocation. Each image was analyzed twice (coefficient of variation for repeated measurements of the same quadriceps scan: 0.45 ± 0.40%), and the average of the two measurements was used for statistical analysis.

In the single exercise bout trial, MVC of the knee extensor muscles of the intervention leg was assessed both isometrically and isokinetically (slow (30°/s) and fast speed (180°/s)) using a Kinetic Communicator dynomometer (Kin-Com, model 500-11, Kinetic Communicator, Chattecx, Chattanooga, TN), as previously described^88^. Rate of force development was assessed from 0-200 ms. Similarly, in the 12-week training trial, MVC of the knee extensor muscles of the right leg was assessed isometrically using a Biodex dynamometer (Biodex Multi-Joint System 4, Software version 4.63, Biodex Medical Systems, Shirley NY, USA). In both sub-studies, participants warmed up on a stationary bike for 5 minutes at a self-selected pace. In participants undergoing the 12-week training intervention, physical function was additionally assessed using the 30-s chair-stand test, 6-min walk test, and timed up-and-go test, performed according to standardized protocols^89–91^

Whole body DXA scans were acquired with a Lunar DPX-IQ scanner (GE-Healthcare) and analyzed in Encore v. 16. Participants were positioned in a standardized manner on the scanner, and regions of interest were placed based on the default definitions provided by the software. Outcomes were lean body mass (LBM), lean leg mass (LLM), and whole-body fat percentage. Bone data from a subset of the HSCT survivors have previously been reported^75^.

### Blood sample

Blood samples were drawn from an antecubital vein and analyzed for hematological parameters, markers of inflammation and muscle injury, metabolic and endocrine function, lipid profile, electrolyte balance, and nutritional and immunological markers, following standard clinical procedures at the Department of Clinical Biochemistry, Bispebjerg Hospital. All blood samples for analyses influenced by dietary intake were obtained in the fasted state (lipid profile). For participants in the single exercise bout trial, blood samples were collected at baseline and seven days after the exercise bout. For participants in the 12-week training intervention, blood samples were collected at baseline and after completion of the training program For serum preparation, blood was allowed to coagulate at room temperature for 15–30 minutes, then centrifuged at 4,000 rpm for 10 minutes. The resulting serum was aliquoted and stored at −80 °C until analysis. Circulating CAF concentrations were measured using the Human Agrin SimpleStep ELISA Kit (ab216945, Abcam, Cambridge, United Kingdom), according to the manufacturer’s instructions. Samples were diluted 1:5 with the provided diluent and analyzed in duplicate. Optical density was measured at 450 nm using a microplate ELISA reader (Multiskan FC, Thermo Scientific). For PBMC isolation, blood was collected in sodium citrate tubes (362781, Biosciences), subjected to density gradient centrifugation, and frozen in fetal bovine serum (FBS) containing 10% dimethyl sulfoxide at −80 °C using a controlled-rate freezing container. Samples were transferred to −130 °C storage the following day.

### Oral-glucose tolerance test

A 3-hour oral glucose tolerance test (OGTT) was performed following an overnight fast. Participants ingested a 75 g glucose solution, and venous blood samples were collected via a peripheral venous catheter in the antecubital vein at regular intervals (−10, −5, +7, +15, +30, +45, +60, +90, +120, +150, and +180 minutes) for assessment of glucose and C-peptide.

### Muscle biopsies

Muscle biopsies were obtained under local anesthesia using a Bergström needle with manual suction^92^. In the single exercise bout trial, biopsies were taken from the middle portion of the vastus lateralis muscle in both the Rested and Exercised legs. In the 12-week training trial, a biopsy was taken from the middle portion of the vastus lateralis muscle before and after the intervention. Pieces of muscle were embedded in Tissue-Tek, rapidly frozen by immersion in isopentane precooled in liquid nitrogen and stored at −80 °C for later analysis. In a subset of participants from the single exercise bout trial, a portion of the tissue was used for *in vitro* experiments (see cell culture section below).

### Single-nucleus RNA sequencing

Free nuclei were isolated from muscle (Rested and Exercised leg) and PBMCs using the Minute Detergent-Free Nuclei Isolation kit (NI-024-INV, MoBiTec, Hamburg, Germany) from four HSCT and four controls. ∼25 mg frozen muscle tissue was cut into small pieces with a scalpel and homogenized in a 2 ml Dounce Tissue Grinder (10749155, Fisher Scientific, Roskilde, Denmark) together with 250 µl Buffer A (from the MoBiTec kit). The homogenate was transferred to the Minute filter and additional 250 µl Buffer A was used to wash the Dounce tube and also transferred to the Minute filter. ∼2 million PBMCs were thawed and spun at 300 g for 5 min at 4°C. The pellet was resuspended in 500 µl Buffer A. Even though PBMCs were already single cells, they were processed for nuclei like the muscle to enable comparison with the immune cells inside the muscle. Both muscle and PBMC suspensions were incubated for 10 min on ice and spun through the filters at 16,000 g in 20 sec. at 4°C to release the nuclei. The pellets were resuspended by short vortex, transferred to a new filter and spun again. The pellets were resuspended again by vortex for 10 sec and the nuclei spun down at 500 g for 3 min at 4°C. The supernatants were removed, and the nuclei pellets resuspended in 300 µl Buffer B by short vortex. Additional 500 µl Buffer B was pipetted below the suspension and the tubes spun at 600 g for 10 min at 4°C. The supernatants were carefully removed, and the nuclei resuspended in 100 µl Anti-clumping buffer by vortex for 10 min. 10 µl of the suspensions were mixed with 10 µl ViaStain AOPI staining solution (13366169, Fisher Scientific) and the nuclei manually counted in a Neubauer hemocytometer. The total yield was 32-231 thousand nuclei for muscle and 8-126 thousand for PBMCs. 8-23 thousand estimated nuclei per sample were used to generate a snRNAseq library using the Chromium Next GEM Single Cell 3′ Kit v3.1 (PN1000269; 10X Genomics, Pleasanton, CA) according to the manufacturer’s protocol. The snRNAseq libraries were sequenced on an Illumina NovaSeq 6000 machine, obtaining 700-1300 million read pairs with 2×150 bp sequencing, by GENEWIZ (Leipzig, Germany).

### Single-nucleus RNA sequencing bioinformatics

Sequencing files were demultiplexed, barcodes were processed and mapped to the human genome using GENCODE Release 38 (GRCh38.p13), and reads were counted using Cell Ranger v7.0.0 (10X Genomics). Only uniquely mapped reads were used. The presence of ambient RNA was identified by examining the frequency distribution of total UMI counts and partially corrected for using CellBender v0.2.2^93^, which was also used to identify nuclei containing droplets. Scrublet vDec2020, which calculates a doublet score based on the observed and synthetic transcriptomes in principal component analysis (PCA) space^94^, was run three times on each data set to identify and remove doublets (2-8% per sample). The nuclei were further filtered for low complexity (< 100 genes) and high mitochondrial content (>2% of transcripts). Souporcell v2.5 was used to identify donor versus recipient nuclei^95^. For each HSCT subject, paired PBMC and muscle samples were analyzed together using Souporcell (options -k 2 --min_alt 10 --min_ref 10 --max_loci 2048), leveraging the European population allele frequencies (minor allele frequency > 0.01) derived from the 1000 Genomes Project^96^. Souporcell was able to classify 99.9% of the nuclei. The 87 unclassified nuclei were removed. The final data set contains 180,419 (6,888-18,956) nuclei from muscle and 118,271 (6,284-25,639) nuclei from PBMC.

Seurat v5.1.0 was used for analysis of the cleaned data sets^97^. Data sets were normalized with SCTransform and integrated using Seurat IntegrateLayers (dims = 1:30, method = RPCAIntegration). PBMC samples were integrated and further analyzed separately from the muscle samples. For the main analysis, clustering was performed using 30 principal components (PCs) on integrated data at a resolution of 0.3. This resulted in 15 clusters for muscle and 20 clusters for PBMC. The clusters were given names based on the expression of known marker genes. To fine-tune the clustering, the clusters that belonged to a broader visual cluster (e.g., different myonuclei clusters) were grouped together and reclustered at a resolution of 0.3, resulting in 63 and 45 subclusters for muscle and PBMC, respectively.

Differences in nuclei abundance (Exercise effects by comparing the two legs) was tested using Wald test in DESeq2 (nNuclei ∼Treatment + Subject, test = “Wald”)^98^. Each cell type was tested separately. For differential expression, DESeq2 Likelihood Ratio Test was performed using pseudobulk expression (total counts for cell type within each sample). For differences between HSCT and controls, the model was: Transcript ∼ Group, test=“LRT”, reduced=∼1. For general exercise effect in muscle the model was: Transcript ∼ Subject + Treatment, test = “LRT”, reduced=∼Subject. For HSCT influence on the Exercise effect in muscle the interaction between Group (HSCT/Control) and Treatment (Exercise/Rest) was tested: Transcript ∼ Group + Group:SubjGroupNum + Group:Treatment, test =“LRT”, reduced=∼Group + Group:SubjGroupNum. SubjGroupNum is Subject ID number within each group to avoid ”Model matrix not full rank” error as described in the DESeq2 vignette. To assess markers for the individual cell clusters each cell cluster was compared to each of the other cell clusters using the pseudobulk expression: Transcript ∼ Subject + CellType, test = “LRT”, reduced=∼Subject. All log2FoldChanges shown are shrunken using lfcshrink from the DESeq2 package. Gene ontology (GO) analysis for biological pathways was performed using topGO v2.56.0 (classic algorithm, Fisher statistic; Bioconductor).

### Cell Culture and Co-culture Experiments

Mononucleated muscle cells were isolated from skeletal muscle biopsies as previously described^61^. Briefly, tissue samples were transferred to a sterile environment, trimmed of visible fat and connective tissue, and subjected to mechanical and enzymatic digestion to release single cells. The resulting cell suspension was expanded for 6–10 days and sorted using CD56 magnetic beads into a CD56⁺ fraction (enriched for MuSCs) and a CD56⁻ fraction (predominantly fibroblasts). Aliquots were frozen in fetal bovine serum (FBS) supplemented with 10% dimethyl sulfoxide (DMSO) at −80 °C using a controlled-rate freezing container and subsequently transferred to liquid nitrogen for long-term storage. Upon initiation of experiments, cells were rapidly thawed, allowed to expand for 4 days in flasks, and sorted a second time to ensure purity. Sorted MuSCs (viability 97 ± 2%) were seeded at 10,000 cells/cm² in growth medium on glass coverslips in 24-well plates for one day. At this point, the medium was replaced with differentiation medium to induce fusion over the following 3 days, and cells were harvested for immunofluorescence and qPCR analyses. Immunofluorescence staining was performed using desmin (AB32362; Abcam) and myogenin (F5D; DSHB). Images were acquired using a 10×/0.30 NA objective with a 0.5× camera (DP71, Olympus) mounted on a BX51 Olympus microscope. Four images were obtained from predefined positions (north, south, east, and west of the center) for each sample. A minimum of 600 nuclei were counted per sample. Fusion was quantified as the number of nuclei within myotubes containing ≥3 nuclei, expressed relative to the total number of desmin-positive nuclei. The RNA was purified using TriReagent as previously described^61^. 60 ng total RNA was converted to cDNA using qScript cDNA SuperMix (Qiagen, Beverly, MA, USA) in 20 µl. 0.25 µl cDNA was amplified with Quantitect SYBR Green PCR mix (Qiagen) and 100 nM primers in 15 µl total volume on a CFX96 (Bio-Rad, Hercules, Ca. USA). From RNAseq analysis of human myoblast cell cultures^61^, we observed two different splice forms of XRRA1, involving exons 1-4, with and without exon 3. Therefore, four different primer sets were tested, covering the region (Table S5). Oligonucleotides corresponding to the expected PCR were used for standard curve and melting point verification. The data were normalized to RPLP0 mRNA.

For co-culture experiments, as above for the myogenic fusion assay above, thawed MuSCs were expanded for 4 days, sorted a second time, and then plated in growth medium at a density of 10,000 cells/cm^2^ on glass coverslips placed in 24-well plates. After 1 day, medium was switched to differentiation medium for one more day, after which motor neurons were added. Primary motor neurons were isolated from E15 Sprague Dawley rat embryos as previously described^61^. Briefly, spinal cords were dissected, mechanically and enzymatically dissociated, and subjected to density gradient centrifugation to enrich for motor neurons. Across four (rat) experiments, motor neurons were plated at 5000 cells/cm^2^ directly onto the MuSCs, in neurobasal medium (replacing 50% of the differentiation medium), and maintained for 2 days. In parallel, and across two (rat) experiments, MuSC-conditioned medium, collected from the MuSC-only cultures at the end of the fusion phase, was added at 50% concentration to motor neuron mono-cultures (plated at 5000 cells/cm^2^ density in 24-well plates containing glass coverslips and Poly-L-Ornithine (P8638; Sigma-Aldrich) and Laminin (L2020; Sigma-Aldrich) coating, and incubated for 2 days to assess paracrine effects on neuronal viability. Motor neuron survival was assessed by immunofluorescence for ChAT⁺ and Tau1⁺ markers (Table S6). For both co-culture and conditioned medium experiments, cells were imaged using a 10×/0.30 NA objective with a 0.5× camera (DP71, Olympus) mounted on a BX51 Olympus microscope. Four images were obtained from predefined positions (north, south, east, and west of the center) for each sample. To account for batch-to-batch variation in motor neuron isolation experiments, HSCT and healthy control cells/conditioned medium were included in each experiment batch, and neuron counts per imaged area were normalized to the mean value of the respective experiment. The RNA was purified as above, but with the addition of 400 ng yeast total RNA per sample as previously described^61^, in order to minimize loss of rat RNA due to low amount. Half of the purified total RNA (5 ul) was used for cDNA synthesis and qPCR as described above but with rat specific primers (Table S5).

### Muscle histology analyses

10 µm-thick muscle biopsy sections were prepared for histological analyses. Immunofluorescence staining was used to assess the following parameters: Denervated fibers, fiber type grouping, fiber type–specific size and shape and myonuclei. Details of the primary and secondary antibodies are provided in Table S6. For assessment of denervated fibers, sections were stained for NCAM (AB5032; Sigma-Aldrich), dystrophin (D8168; Sigma-Aldrich), and MyHC I (A4.951; DSHB). Denervated fibers were quantified relative to total fiber number as previously described in detail^88^. From the same sections, fiber type grouping was assessed by quantifying the proportion of grouped type I (MyHC I–positive) and type II (MyHC I–negative) fibers according to the method of Jennekens, with minor modifications^99^. Specifically, fibers fully enclosed by fibers of the same type, together with the enclosing fibers, were counted, and the grouped fiber counts were adjusted for the overall proportion of type I and type II fibers in the sample. For fiber type–specific morphology and fiber type distribution, sections were stained with MyHC I (BA.D5; DSHB) and laminin (L9393; Sigma-Aldrich). Cross-sectional area (CSA), shape factor index (SFI), and fiber type distribution were determined as previously described^30^. Myonuclear content was assessed in sections stained with PCM1 (HPA023370; Sigma-Aldrich), MyHC I (BA.D5; DSHB), and collagen IV (AB769; Millipore). For XRRA1 staining, sections were incubated with the primary anti-XRRA1 antibody alongside matched negative controls in which the primary antibody was omitted. After secondary antibody incubation, images were acquired under identical settings for all samples. XRRA1 expression was quantified as the area of XRRA1-positive pixels, with background signal estimated from negative-control sections and subtracted from all measurements. Samples were imaged using either a 10×/0.30 NA objective with a 0.5× camera (DP71, Olympus) mounted on a BX51 Olympus microscope, or a 20×/0.80 NA objective on an LSM710 Zeiss confocal microscope for the PCM1 staining.

### Exercise interventions

In the single bout exercise trial, participants performed a single bout of unilateral heavy resistance exercise in a dynamometer (KinCom, Chattanooga Group, TN, USA). The protocol was conducted on one leg (designated as the Exercise leg), while the contralateral leg served as the rested Control leg. The exercise session consisted of two rounds separated by a 5–10-minute rest period. Each round included four sets of 10 concentric contractions performed at an angular velocity of 30°/s, aiming at an intensity greater than 70% of the participant’s MVC. This was followed by five sets of five eccentric contractions at the same angular velocity but aiming at an intensity exceeding 100% of MVC. In the second round, the contraction velocity was increased to 180°/s. Verbal encouragement and real-time visual feedback were provided throughout the session to maximize effort. Participants rested for 1.5–2.5 minutes between sets. Ratings of perceived exertion (RPE) were recorded after each set. Force output was measured during the first, middle, and last repetition of every set to evaluate fatigue and performance consistency. Isometric MVC was reassessed two minutes after the final exercise set.

In the training trial, participants completed 36 (range: 34-38) supervised sessions of heavy resistance training (HReT) over a 12-week period, corresponding to three sessions per week. Each session began with a standardized warm-up on a stationary bicycle, followed by a structured resistance training program targeting both lower and upper body muscle groups. The lower body exercises included horizontal leg press, seated leg extension, and seated leg curl. Two upper body exercises - pulldown and shoulder press - were also performed. One-repetition maximum (1RM) was assessed in the lower body exercises five times during the intervention (prior to sessions 1, 8, 18, 28, and 36), and the resulting values were used to individualize training loads for subsequent sessions. The program followed a progressive resistance model, with intensity increasing from approximately a 15-repetition maximum (RM) load at the start to a 6-RM load by the end of the intervention. All training sessions were documented in individual training diaries, including the number of sets, repetitions, and load used, as well as machine settings. An overview of the full training program is presented in Table S4.

### Statistical analyses

Statistical analyses were performed using GraphPad Prism (version 10.6; GraphPad Software). Data are presented as mean ± standard deviation (SD) with individual data points, or as median with individual data points for variables not meeting normality assumptions. PCR data are shown as geometric means with standard error of the mean (SEM), and were log-transformed before statistical testing. Baseline group comparisons were conducted using unpaired two-tailed t-tests for normally distributed variables and Mann–Whitney U tests for non-normally distributed variables, as indicated in respective figure legends. Where an effect of sex was visually apparent, a two-way mixed-effects model with group and sex as fixed factors was applied, followed by Fisher’s least significant difference (LSD) post hoc testing where appropriate. For XRRA1 qPCR analyses, group differences were assessed using a three-way mixed effects model with group, splice form, and cell condition as fixed factors. Longitudinal changes over time were analyzed using two-way repeated-measures mixed-effects models with group and time as fixed factors, followed by Fisher’s LSD post hoc testing where appropriate. Statistical significance was accepted at p < 0.05. Trends are reported where relevant.

## Resource availability

### Lead contact

Further information or requests for data and resources should be directed to, and will be fulfilled by, the lead contact, Abigail L. Mackey

### Materials availability

This study did not generate new unique reagents.

### Data and code availability

The snRNAseq data have been deposited in Array-Express with the data set identifier E-MTAB-16474 (muscle) and E-MTAB-16475 (PBMC).

All other data are provided in Excel files.

## Supporting information

STROBE checklist

## Data Availability

All data produced in the present work are contained in the manuscript

## Acknowledgement

This study was supported by funding from the Lundbeck Foundation (R402-2022-1387 to CS, R344-2020–254 and R485-2025-192 to ALM), The Danish Childhood Cancer Foundation (2017-2021, 2020-6792 to KM), The Research Foundation of Rigshospitalet (Rigshospitalets Forskningsfond), Copenhagen University Hospital (to AN), Danish Cancer Research Foundation (to AN), The Svend Andersen Foundation (to KM).

The monoclonal antibodies BA.D5 (type I MyHC), developed by Schiaffino, S., PAX7, developed by Kawakami, A., MYOG (myogenin), developed by Wright, W.E., and A4.951 (type I MyHC), developed by Blau, H.M., were obtained from the Developmental Studies Hybridoma Bank (DSHB), created by the NICHD of the NIH and maintained at The University of Iowa, Department of Biology, Iowa City, IA, USA.

The authors sincerely thank all study participants for their time, commitment, and enthusiasm, as well as the students involved in data collection. We warmly acknowledge laboratory technician Anja Jokipii-Utzon for assistance with qPCR analyses and preparation of samples for RNA sequencing, and laboratory technician Ann-Christina Ronnié Reimann for technical assistance with muscle biopsies and for performing ELISA analyses. We thank René B. Svensson (Institute of Sports Medicine Copenhagen, Bispebjerg-Frederiksberg Hospital, and Center for Fast Ultrasound Imaging, Department of Health Technology, Technical University of Denmark, Kongens Lyngby, Denmark) for assistance with magnetic resonance imaging analysis.

We thank the Core Facility for Flow Cytometry and Single Cell Analysis, Faculty of Health and Medical Sciences, University of Copenhagen, for technical support with fluorescence-activated cell sorting (FACS) and 10x Genomics single-cell sequencing. We also acknowledge the Core Facility for Integrated Microscopy, Faculty of Health and Medical Sciences, University of Copenhagen, for access to slide-scanning and imaging equipment. We further acknowledge the Department of Clinical Biochemistry, Faculty of Health Sciences, Bispebjerg Hospital, University of Copenhagen, for access to animal and cell culture facilities. Special thanks are extended to Jens Hannibal and the animal caretakers for expert assistance with animal handling and care.

## Author contributions

Conceptualization: C.S., A.N., M.K., K.M., A.L.M.

Data curation: C.S., A.N., L.M.K., P.S., V.D.S., J.L.A., K.H.M.

Formal analysis: C.S., A.N., P.S., V.D.S.

Funding acquisition: C.S., A.N., K.M., A.L.M., M.K.

Investigation: C.S., A.N., L.M.K., P.S., J.G., M.K., K.H.M.

Methodology: C.S., A.N., M.K.F., P.S., M.K., J.L.A., K.M., A.L.M.

Project administration: C.S., A.N., K.M., A.L.M., L.M.K.

Resources: M.K., K.M., A.L.M.

Supervision: K.M., A.L.M.

Visualization: C.S., P.S. A.L.M.

Writing – original draft: C.S., A.L.M.

Writing – review & editing: All authors.

## Supplemental information titles and legends

Document S1. Tables S1–S6 and Figures S1–S7

## References

1. Passweg, J.R., Baldomero, H., Chabannon, C., Basak, G.W., de la Cámara, R., Corbacioglu, S., Dolstra, H., Duarte, R., Glass, B., Greco, R., et al. (2021). Hematopoietic cell transplantation and cellular therapy survey of the EBMT: monitoring of activities and trends over 30 years. Bone Marrow Transplant 56, 1651–1664. 10.1038/s41409-021-01227-8.

2. Kliman, D., Nivison-Smith, I., Gottlieb, D., Hamad, N., Kerridge, I., Purtill, D., Szer, J., and Ma, D. (2020). Hematopoietic Stem Cell Transplant Recipients Surviving at Least 2 Years from Transplant Have Survival Rates Approaching Population Levels in the Modern Era of Transplantation. Biol Blood Marrow Transplant 26, 1711–1718. 10.1016/j.bbmt.2020.03.005.

3. Baker, K.S., Ness, K.K., Steinberger, J., Carter, A., Francisco, L., Burns, L.J., Sklar, C., Forman, S., Weisdorf, D., Gurney, J.G., et al. (2007). Diabetes, hypertension, and cardiovascular events in survivors of hematopoietic cell transplantation: a report from the bone marrow transplantation survivor study. Blood 109, 1765–1772. 10.1182/blood-2006-05-022335.

4. Arora, M., Sun, C.-L., Ness, K.K., Teh, J.B., Wu, J., Francisco, L., Armenian, S.H., Schad, A., Namdar, G., Bosworth, A., et al. (2016). Physiologic Frailty in Nonelderly Hematopoietic Cell Transplantation Patients: Results From the Bone Marrow Transplant Survivor Study. JAMA Oncol 2, 1277–1286. 10.1001/jamaoncol.2016.0855.

5. Lorenc, A., Hamilton-Shield, J., Perry, R., Stevens, M., and CTYA HSCT Adipose and Muscle Late Effects Working Group (2020). Body composition after allogeneic haematopoietic cell transplantation/total body irradiation in children and young people: a restricted systematic review. J Cancer Surviv 14, 624–642. 10.1007/s11764-020-00871-1.

6. Mejdahl Nielsen, M., Mathiesen, S., Suominen, A., Sørensen, K., Ifversen, M., Mølgaard, C., Lähteenmäki, P.M., Juul, A., Jahnukainen, K., and Müller, K. (2021). Altered body composition in male long-term survivors of paediatric allogeneic haematopoietic stem cell transplantation: impact of conditioning regimen, chronic graft-versus-host disease and hypogonadism. Bone Marrow Transplant 56, 457–460. 10.1038/s41409-020-01038-3.

7. Suominen, A., Haavisto, A., Mathiesen, S., Mejdahl Nielsen, M., Lähteenmäki, P.M., Sørensen, K., Ifversen, M., Mølgaard, C., Juul, A., Müller, K., et al. (2022). Physical Fitness and Frailty in Males after Allogeneic Hematopoietic Stem Cell Transplantation in Childhood: A Long-Term Follow-Up Study. Cancers (Basel) 14, 3310. 10.3390/cancers14143310.

8. Koeppen, S., Thirugnanasambanthan, A., and Koldehoff, M. (2014). Neuromuscular complications after hematopoietic stem cell transplantation. Support Care Cancer 22, 2337–2341. 10.1007/s00520-014-2225-0.

9. Fridh, M.K., Simonsen, C., Schmidt-Andersen, P., Nissen, A.A., Christensen, J.F., Larsen, A., Mackey, A.L., Larsen, H.B., and Müller, K. (2021). Cardiorespiratory fitness and physical performance after childhood hematopoietic stem cell transplantation: a systematic review and meta-analysis. Bone Marrow Transplant. 10.1038/s41409-021-01370-2.

10. Jarden, M., Baadsgaard, M.T., Hovgaard, D.J., Boesen, E., and Adamsen, L. (2009). A randomized trial on the effect of a multimodal intervention on physical capacity, functional performance and quality of life in adult patients undergoing allogeneic SCT. Bone Marrow Transplant 43, 725–737. 10.1038/bmt.2009.27.

11. San Juan, A.F., Fleck, S.J., Chamorro-Viña, C., Maté-Muñoz, J.L., Moral, S., Pérez, M., Cardona, C., Del Valle, M.F., Hernández, M., Ramírez, M., et al. (2007). Effects of an intrahospital exercise program intervention for children with leukemia. Med Sci Sports Exerc 39, 13–21. 10.1249/01.mss.0000240326.54147.fc.

12. Chamorro-Viña, C., Ruiz, J.R., Santana-Sosa, E., González Vicent, M., Madero, L., Pérez, M., Fleck, S.J., Pérez, A., Ramírez, M., and Lucía, A. (2010). Exercise during hematopoietic stem cell transplant hospitalization in children. Med Sci Sports Exerc 42, 1045–1053. 10.1249/MSS.0b013e3181c4dac1.

13. Rosenhagen, A., Bernhörster, M., Vogt, L., Weiss, B., Senn, A., Arndt, S., Siegler, K., Jung, M., Bader, P., and Banzer, W. (2011). Implementation of structured physical activity in the pediatric stem cell transplantation. Klin Padiatr 223, 147–151. 10.1055/s-0031-1271782.

14. Davis, N.L., Tolfrey, K., Jenney, M., Elson, R., Stewart, C., Moss, A.D., Cornish, J.M., Stevens, M.C.G., and Crowne, E.C. (2020). Combined resistance and aerobic exercise intervention improves fitness, insulin resistance and quality of life in survivors of childhood haemopoietic stem cell transplantation with total body irradiation. Pediatr Blood Cancer 67, e28687. 10.1002/pbc.28687.

15. Janssen, I., Heymsfield, S.B., and Ross, R. (2002). Low relative skeletal muscle mass (sarcopenia) in older persons is associated with functional impairment and physical disability. J Am Geriatr Soc 50. 10.1046/j.1532-5415.2002.50216.x.

16. Kalyani, R.R., Corriere, M., and Ferrucci, L. (2014). Age-related and disease-related muscle loss: the effect of diabetes, obesity, and other diseases. Lancet Diabetes Endocrinol 2, 819–829. 10.1016/S2213-8587(14)70034-8.

17. Abramowitz, M.K., Hall, C.B., Amodu, A., Sharma, D., Androga, L., and Hawkins, M. (2018). Muscle mass, BMI, and mortality among adults in the United States: A population-based cohort study. PLoS One 13, e0194697. 10.1371/journal.pone.0194697.

18. Roberts, M.D., McCarthy, J.J., Hornberger, T.A., Phillips, S.M., Mackey, A.L., Nader, G.A., Boppart, M.D., Kavazis, A.N., Reidy, P.T., Ogasawara, R., et al. (2023). Mechanisms of mechanical overload-induced skeletal muscle hypertrophy: current understanding and future directions. Physiol Rev 103, 2679–2757. 10.1152/physrev.00039.2022.

19. Zhu, W.G., Thomas, A.C.Q., Wilson, G.M., McGlory, C., Hibbert, J.E., Flynn, C.G., Sayed, R.K.A., Paez, H.G., Meinhold, M., Jorgenson, K.W., et al. (2025). Identification of a resistance-exercise-specific signalling pathway that drives skeletal muscle growth. Nat Metab 7, 1404–1423. 10.1038/s42255-025-01298-7.

20. Schiaffino, S., Bormioli, S.P., and Aloisi, M. (1976). The fate of newly formed satellite cells during compensatory muscle hypertrophy. Virchows Arch B Cell Pathol 21, 113–118. 10.1007/BF02899148.

21. Murach, K.A., Fry, C.S., Dupont-Versteegden, E.E., McCarthy, J.J., and Peterson, C.A. (2021). Fusion and beyond: Satellite cell contributions to loading-induced skeletal muscle adaptation. FASEB J 35, e21893. 10.1096/fj.202101096R.

22. Epelman, S., Lavine, K.J., and Randolph, G.J. (2014). Origin and functions of tissue macrophages. Immunity 41, 21–35. 10.1016/j.immuni.2014.06.013.

23. Paulsen, G., Crameri, R., Benestad, H.B., Fjeld, J.G., Mørkrid, L., Hallén, J., and Raastad, T. (2010). Time course of leukocyte accumulation in human muscle after eccentric exercise. Med Sci Sports Exerc 42, 75–85. 10.1249/MSS.0b013e3181ac7adb.

24. Brazelton, T.R., Nystrom, M., and Blau, H.M. (2003). Significant differences among skeletal muscles in the incorporation of bone marrow-derived cells. Dev Biol 262, 64–74. 10.1016/s0012-1606(03)00357-9.

25. LaBarge, M.A., and Blau, H.M. (2002). Biological progression from adult bone marrow to mononucleate muscle stem cell to multinucleate muscle fiber in response to injury. Cell 111, 589–601. 10.1016/s0092-8674(02)01078-4.

26. Sherwood, R.I., Christensen, J.L., Weissman, I.L., and Wagers, A.J. (2004). Determinants of skeletal muscle contributions from circulating cells, bone marrow cells, and hematopoietic stem cells. Stem Cells 22, 1292–1304. 10.1634/stemcells.2004-0090.

27. Palermo, A.T., LaBarge, M.A., Doyonnas, R., Pomerantz, J., and Blau, H.M. (2005). Bone marrow contribution to skeletal muscle: A physiological response to stress. Developmental Biology 279, 336–344. 10.1016/j.ydbio.2004.12.024.

28. Jiang, S., Walker, L., Afentoulis, M., Anderson, D.A., Jauron-Mills, L., Corless, C.L., and Fleming, W.H. (2004). Transplanted human bone marrow contributes to vascular endothelium. Proc Natl Acad Sci U S A 101, 16891–16896. 10.1073/pnas.0404398101.

29. Strömberg, A., Jansson, M., Fischer, H., Rullman, E., Hägglund, H., and Gustafsson, T. (2013). Bone marrow derived cells in adult skeletal muscle tissue in humans. Skelet Muscle 3, 12. 10.1186/2044-5040-3-12.

30. Soendenbroe, C., Karlsen, A., Svensson, R.B., Kjaer, M., Andersen, J.L., and Mackey, A.L. (2024). Marked irregular myofiber shape is a hallmark of human skeletal muscle ageing and is reversed by heavy resistance training. J Cachexia Sarcopenia Muscle 15, 306–318. 10.1002/jcsm.13405.

31. Hepple, R.T., Mackinnon, S.L., Thomas, S.G., Goodman, J.M., and Plyley, M.J. (1997). Quantitating the capillary supply and the response to resistance training in older men. Pflugers Arch 433, 238–244. 10.1007/s004240050273.

32. McGraw, J.M., Thelen, F., Hampton, E.N., Bruno, N.E., Young, T.S., Havran, W.L., and Witherden, D.A. (2021). JAML promotes CD8 and γδ T cell antitumor immunity and is a novel target for cancer immunotherapy. J Exp Med 218, e20202644. 10.1084/jem.20202644.

33. Flynn, R., Allen, J.L., Luznik, L., MacDonald, K.P., Paz, K., Alexander, K.A., Vulic, A., Du, J., Panoskaltsis-Mortari, A., Taylor, P.A., et al. (2015). Targeting Syk-activated B cells in murine and human chronic graft-versus-host disease. Blood 125, 4085–4094. 10.1182/blood-2014-08-595470.

34. Kedlian, V.R., Wang, Y., Liu, T., Chen, X., Bolt, L., Tudor, C., Shen, Z., Fasouli, E.S., Prigmore, E., Kleshchevnikov, V., et al. (2024). Human skeletal muscle aging atlas. Nat Aging. 10.1038/s43587-024-00613-3.

35. Soendenbroe, C., Andersen, J.L., and Mackey, A.L. (2021). Muscle-nerve communication and the molecular assessment of human skeletal muscle denervation with aging. American Journal of Physiology-Cell Physiology 321, C317–C329. 10.1152/ajpcell.00174.2021.

36. Ham, A.S., Lin, S., Tse, A., Thürkauf, M., McGowan, T.J., Jörin, L., Oliveri, F., and Rüegg, M.A. (2025). Single-nuclei sequencing of skeletal muscle reveals subsynaptic-specific transcripts involved in neuromuscular junction maintenance. Nat Commun 16, 2220. 10.1038/s41467-025-57487-1.

37. Perez, K., Ciotlos, S., McGirr, J., Limbad, C., Doi, R., Nederveen, J.P., Nilsson, M.I., Winer, D.A., Evans, W., Tarnopolsky, M., et al. (2022). Single nuclei profiling identifies cell specific markers of skeletal muscle aging, frailty, and senescence. Aging (Albany NY) 14, 9393–9422. 10.18632/aging.204435.

38. Nieves-Rodriguez, S., Barthélémy, F., Woods, J.D., Douine, E.D., Wang, R.T., Scripture-Adams, D.D., Chesmore, K.N., Galasso, F., Miceli, M.C., and Nelson, S.F. (2023). Transcriptomic analysis of paired healthy human skeletal muscles to identify modulators of disease severity in DMD. Front Genet 14, 1216066. 10.3389/fgene.2023.1216066.

39. Hansen, M., Grothen, J.E.R., Karlsen, A., Martinez, J.M., Sidiropoulos, N., Helge, J.W., Pedersen, T.Å., and Dela, F. (2025). The skeletal muscle response to high-intensity training assessed by single-nucleus RNA-sequencing is blunted in individuals with type 2 diabetes. J Physiol. 10.1113/JP288368.

40. Karlsen, A., Yeung, C.-Y.C., Schjerling, P., Denz, L., Hoegsbjerg, C., Jakobsen, J.R., Krogsgaard, M.R., Koch, M., Schiaffino, S., Kjaer, M., et al. (2023). Distinct myofibre domains of the human myotendinous junction revealed by single nucleus RNA-seq. J Cell Sci, jcs.260913. 10.1242/jcs.260913.

41. Soule, T.G., Pontifex, C.S., Rosin, N., Joel, M.M., Lee, S., Nguyen, M.D., Chhibber, S., and Pfeffer, G. (2023). A protocol for single nucleus RNA-seq from frozen skeletal muscle. Life Sci Alliance 6, e202201806. 10.26508/lsa.202201806.

42. Rubenstein, A.B., Smith, G.R., Zhang, Z., Chen, X., Chambers, T.L., Ruf-Zamojski, F., Mendelev, N., Cheng, W.S., Zamojski, M., Amper, M.A.S., et al. (2025). Integrated single-cell multiome analysis reveals muscle fiber-type gene regulatory circuitry modulated by endurance exercise. Genome Res, gr.280051.124. 10.1101/gr.280051.124.

43. Lovrić, A., Rassolie, A., Alam, S., Mandić, M., Saini, A., Altun, M., Fernandez-Gonzalo, R., Gustafsson, T., and Rullman, E. (2022). Single-cell sequencing deconvolutes cellular responses to exercise in human skeletal muscle. Commun Biol 5, 1121. 10.1038/s42003-022-04088-z.

44. Pass, C.G., Palzkill, V., Tan, J., Kim, K., Thome, T., Yang, Q., Fazzone, B., Robinson, S.T., O’Malley, K.A., Yue, F., et al. (2023). Single-Nuclei RNA-Sequencing of the Gastrocnemius Muscle in Peripheral Artery Disease. Circ Res 133, 791–809. 10.1161/CIRCRESAHA.123.323161.

45. Lai, Y., Ramírez-Pardo, I., Isern, J., An, J., Perdiguero, E., Serrano, A.L., Li, J., García-Domínguez, E., Segalés, J., Guo, P., et al. (2024). Multimodal cell atlas of the ageing human skeletal muscle. Nature 629, 154–164. 10.1038/s41586-024-07348-6.

46. Eraslan, G., Drokhlyansky, E., Anand, S., Fiskin, E., Subramanian, A., Slyper, M., Wang, J., Van Wittenberghe, N., Rouhana, J.M., Waldman, J., et al. (2022). Single-nucleus cross-tissue molecular reference maps toward understanding disease gene function. Science 376, eabzl4290. 10.1126/science.abl4290.

47. De Micheli, A.J., Spector, J.A., Elemento, O., and Cosgrove, B.D. (2020). A reference single-cell transcriptomic atlas of human skeletal muscle tissue reveals bifurcated muscle stem cell populations. Skelet Muscle 10, 19. 10.1186/s13395-020-00236-3.

48. Rubenstein, A.B., Smith, G.R., Raue, U., Begue, G., Minchev, K., Ruf-Zamojski, F., Nair, V.D., Wang, X., Zhou, L., Zaslavsky, E., et al. (2020). Single-cell transcriptional profiles in human skeletal muscle. Sci Rep 10, 229. 10.1038/s41598-019-57110-6.

49. Nazarian, J., Berry, D.L., Sanjari, S., Razvi, M., Brown, K., Hathout, Y., Vertes, A., Dadgar, S., and Hoffman, E.P. (2011). Evolution and comparative genomics of subcellular specializations: EST sequencing of Torpedo electric organ. Mar Genomics 4, 33–40. 10.1016/j.margen.2010.12.004.

50. Mesak, F.M., Osada, N., Hashimoto, K., Liu, Q.Y., and Ng, C.E. (2003). Molecular cloning, genomic characterization and over-expression of a novel gene, XRRA1, identified from human colorectal cancer cell HCT116Clone2_XRR and macaque testis. BMC Genomics 4, 32. 10.1186/1471-2164-4-32.

51. Nojiri, K., Iwakawa, M., Ichikawa, Y., Imadome, K., Sakai, M., Nakawatari, M., Ishikawa, K.-I., Ishikawa, A., Togo, S., Tsujii, H., et al. (2009). The proangiogenic factor ephrin-A1 is up-regulated in radioresistant murine tumor by irradiation. Exp Biol Med (Maywood) 234, 112–122. 10.3181/0806-RM-189.

52. Collao, N., Johannsen, E.B., Just, J., and De Lisio, M. (2025). Single-cell transcriptomic analysis reveals alterations to cellular dynamics and paracrine signalling in radiation-induced muscle pathology. Am J Physiol Cell Physiol. 10.1152/ajpcell.00115.2025.

53. Singh, V., Singh, R., and Kushwaha, R. (2024). Exploring novel protein biomarkers for early-stage diagnosis and prognosis of T-acute lymphoblastic leukemia (T-ALL). Hematol Transfus Cell Ther 46 *Suppl 6*, S93–S111. 10.1016/j.htct.2024.02.016.

54. Bechshøft, C.J.L., Jensen, S.M., Schjerling, P., Andersen, J.L., Svensson, R.B., Eriksen, C.S., Mkumbuzi, N.S., Kjaer, M., and Mackey, A.L. (2019). Age and prior exercise in vivo determine the subsequent in vitro molecular profile of myoblasts and nonmyogenic cells derived from human skeletal muscle. Am. J. Physiol., Cell Physiol. 316, C898–C912. 10.1152/ajpcell.00049.2019.

55. Liu, M., Hu, P., Tang, B., Yang, Q., Xiang, R., Liu, Y., Li, J., Wu, B., Wu, H., Tian, B., et al. (2024). Endoplasmic reticulum stress-MMRN1 positive feedback contributes to cisplatin resistance in small cell lung cancer. J Thorac Dis 16, 8363–8378. 10.21037/jtd-24-1477.

56. Wen, D., Yan, R., Zhang, L., Zhang, H., Chen, X., and Zhou, J. (2025). Screening of necroptosis-related genes and evaluating the prognostic capacity, clinical value, and the effect of their copy number variations in acute myeloid leukemia. BMC Cancer 25, 71. 10.1186/s12885-025-13439-y.

57. Yun, D., Yang, J.-H., Yang, S., Sim, J.-A., Kim, M., Park, J.W., Jeong, S.Y., Shin, A., Kweon, S.-S., and Song, N. (2025). Novel genetic loci and functional properties of immune-related genes for colorectal cancer survival in Korea. BMC Cancer 25, 456. 10.1186/s12885-025-13819-4.

58. Hepple, R.T., and Rice, C.L. (2016). Innervation and neuromuscular control in ageing skeletal muscle. J. Physiol. (Lond.) 594, 1965–1978. 10.1113/JP270561.

59. Folker, E.S., and Baylies, M.K. (2013). Nuclear positioning in muscle development and disease. Front Physiol 4, 363. 10.3389/fphys.2013.00363.

60. Monti, E., Sarto, F., Sartori, R., Zanchettin, G., Löfler, S., Kern, H., Narici, M.V., and Zampieri, S. (2023). C-terminal agrin fragment as a biomarker of muscle wasting and weakness: a narrative review. J Cachexia Sarcopenia Muscle. 10.1002/jcsm.13189.

61. Soendenbroe, C., Schjerling, P., Bechshøft, C.J.L., Svensson, R.B., Schaeffer, L., Kjaer, M., Chazaud, B., Jacquier, A., and Mackey, A.L. (2025). Muscle fibroblasts and stem cells stimulate motor neurons in an age and exercise-dependent manner. Aging Cell 24, e14413. 10.1111/acel.14413.

62. Fredette, B.J., Miller, J., and Ranscht, B. (1996). Inhibition of motor axon growth by T-cadherin substrata. Development 122, 3163–3171. 10.1242/dev.122.10.3163.

63. Tamáš, M., Pankratova, S., Schjerling, P., Soendenbroe, C., Yeung, C.C., Pennisi, C.P., Jakobsen, J.R., Krogsgaard, M.R., Kjaer, M., and Mackey, A.L. (2021). Mutual stimulatory signaling between human myogenic cells and rat cerebellar neurons. Physiological Reports 9. 10.14814/phy2.15077.

64. Edman, S., Jones Iii, R.G., Jannig, P.R., Fernandez-Gonzalo, R., Norrbom, J., Thomas, N.T., Khadgi, S., Koopmans, P.J., Morena, F., Chambers, T.L., et al. (2024). The 24-hour molecular landscape after exercise in humans reveals MYC is sufficient for muscle growth. EMBO Rep 25, 5810–5837. 10.1038/s44319-024-00299-z.

65. Long, D.E., Peck, B.D., Lavin, K.M., Dungan, C.M., Kosmac, K., Tuggle, S.C., Bamman, M.M., Kern, P.A., and Peterson, C.A. (2022). Skeletal muscle properties show collagen organization and immune cell content are associated with resistance exercise response heterogeneity in older persons. J Appl Physiol (1985) 132, 1432–1447. 10.1152/japplphysiol.00025.2022.

66. Møbjerg, A., Steffen, D., Schjerling, P., Jakobsen, J.R., Jokipii-Utzon, A., Batiuk, M.Y., Khodosevich, K., Krogsgaard, M.R., Izzi, V., Mackey, A.L., et al. (2025). Spatially distinct ECM-producing fibroblasts and myonuclei orchestrate early adaptation to mechanical loading in the human muscle-tendon unit. Am J Physiol Cell Physiol 329, C1775–C1791. 10.1152/ajpcell.00700.2025.

67. Hu, Y.-W., Guo, F.-X., Xu, Y.-J., Li, P., Lu, Z.-F., McVey, D.G., Zheng, L., Wang, Q., Ye, J.H., Kang, C.-M., et al. Long noncoding RNA NEXN-AS1 mitigates atherosclerosis by regulating the actin-binding protein NEXN. J Clin Invest 129, 1115–1128. 10.1172/JCI98230.

68. Porpora, M., Sauchella, S., Rinaldi, L., Delle Donne, R., Sepe, M., Torres-Quesada, O., Intartaglia, D., Garbi, C., Insabato, L., Santoriello, M., et al. (2018). Counterregulation of cAMP-directed kinase activities controls ciliogenesis. Nat Commun 9, 1224. 10.1038/s41467-018-03643-9.

69. Benmerah, A., Briseño-Roa, L., Annereau, J.-P., and Saunier, S. (2023). Repurposing small molecules for nephronophthisis and related renal ciliopathies. Kidney International 104, 245–253. 10.1016/j.kint.2023.04.027.

70. Arnal, J.-F., Lenfant, F., Metivier, R., Flouriot, G., Henrion, D., Adlanmerini, M., Fontaine, C., Gourdy, P., Chambon, P., Katzenellenbogen, B., et al. (2017). Membrane and Nuclear Estrogen Receptor Alpha Actions: From Tissue Specificity to Medical Implications. Physiol Rev 97, 1045–1087. 10.1152/physrev.00024.2016.

71. Morikawa, M., Mitani, Y., Holmborn, K., Kato, T., Koinuma, D., Maruyama, J., Vasilaki, E., Sawada, H., Kobayashi, M., Ozawa, T., et al. (2019). The ALK-1/SMAD/ATOH8 axis attenuates hypoxic responses and protects against the development of pulmonary arterial hypertension. Sci Signal 12, eaay4430. 10.1126/scisignal.aay4430.

72. Seaayfan, E., Nasrah, S., Quell, L., Radi, A., Kleim, M., Schermuly, R.T., Weber, S., Laghmani, K., and Kömhoff, M. (2022). Reciprocal Regulation of MAGED2 and HIF-1α Augments Their Expression under Hypoxia: Role of cAMP and PKA Type II. Cells 11, 3424. 10.3390/cells11213424.

73. Ishida, Y., and Nagata, K. (2011). Hsp47 as a collagen-specific molecular chaperone. Methods Enzymol 499, 167–182. 10.1016/B978-0-12-386471-0.00009-2.

74. Ward, S.K., Wadley, A., Tsai, C. (Anne), Benke, P.J., Emrick, L., Fisher, K., Houck, K.M., Dai, H., Sacoto, M.J.G., Craigen, W., et al. (2024). De novo missense variants in ZBTB47 are associated with developmental delays, hypotonia, seizures, gait abnormalities, and variable movement abnormalities. Am J Med Genet A 194, 17–30. 10.1002/ajmg.a.63399.

75. Krogh, L.M., Nissen, A., Weischendorff, S., Hartmann, B., Andersen, J.L., Holst, J.J., Sørensen, K., Fridh, M.K., Mackey, A.L., and Müller, K. (2024). Bone remodeling in survivors of pediatric hematopoietic stem cell transplantation: Impact of heavy resistance training. Pediatr Blood Cancer, e31159. 10.1002/pbc.31159.

76. Valdez, G., Tapia, J.C., Kang, H., Clemenson, G.D., Gage, F.H., Lichtman, J.W., and Sanes, J.R. (2010). Attenuation of age-related changes in mouse neuromuscular synapses by caloric restriction and exercise. Proc Natl Acad Sci U S A 107, 14863–14868. 10.1073/pnas.1002220107.

77. Paudyal, A., Slevin, M., Maas, H., and Degens, H. (2018). Time course of denervation-induced changes in gastrocnemius muscles of adult and old rats. Exp Gerontol 106, 165–172. 10.1016/j.exger.2018.03.008.

78. Bagley, J.R., Denes, L.T., McCarthy, J.J., Wang, E.T., and Murach, K.A. (2023). The myonuclear domain in adult skeletal muscle fibres: past, present and future. J Physiol 601, 723–741. 10.1113/JP283658.

79. Karlsen, A., Couppé, C., Andersen, J.L., Mikkelsen, U.R., Nielsen, R.H., Magnusson, S.P., Kjaer, M., and Mackey, A.L. (2015). Matters of fiber size and myonuclear domain: Does size matter more than age? Muscle Nerve 52, 1040–1046. 10.1002/mus.24669.

80. Karlsen, A., Bechshøft, R.L., Malmgaard-Clausen, N.M., Andersen, J.L., Schjerling, P., Kjaer, M., and Mackey, A.L. (2019). Lack of muscle fibre hypertrophy, myonuclear addition, and satellite cell pool expansion with resistance training in 83-94-year-old men and women. Acta Physiol (Oxf) 227, e13271. 10.1111/apha.13271.

81. Bachman, J.F., Blanc, R.S., Paris, N.D., Kallenbach, J.G., Johnston, C.J., Hernady, E., Williams, J.P., and Chakkalakal, J.V. (2020). Radiation-Induced Damage to Prepubertal Pax7+ Skeletal Muscle Stem Cells Drives Lifelong Deficits in Myofiber Size and Nuclear Number. iScience 23, 101760. 10.1016/j.isci.2020.101760.

82. Bittner, R.E., Schöfer, C., Weipoltshammer, K., Ivanova, S., Streubel, B., Hauser, E., Freilinger, M., Höger, H., Elbe-Bürger, A., and Wachtler, F. (1999). Recruitment of bone-marrow-derived cells by skeletal and cardiac muscle in adult dystrophic mdx mice. Anat Embryol (Berl) 199, 391–396. 10.1007/s004290050237.

83. Ferrari, G., Cusella-De Angelis, G., Coletta, M., Paolucci, E., Stornaiuolo, A., Cossu, G., and Mavilio, F. (1998). Muscle regeneration by bone marrow-derived myogenic progenitors. Science 279, 1528–1530. 10.1126/science.279.5356.1528.

84. Gussoni, E., Soneoka, Y., Strickland, C.D., Buzney, E.A., Khan, M.K., Flint, A.F., Kunkel, L.M., and Mulligan, R.C. (1999). Dystrophin expression in the mdx mouse restored by stem cell transplantation. Nature 401, 390–394. 10.1038/43919.

85. Ding, J., Adiconis, X., Simmons, S.K., Kowalczyk, M.S., Hession, C.C., Marjanovic, N.D., Hughes, T.K., Wadsworth, M.H., Burks, T., Nguyen, L.T., et al. (2020). Systematic comparison of single-cell and single-nucleus RNA-sequencing methods. Nat Biotechnol 38, 737–746. 10.1038/s41587-020-0465-8.

86. von Elm, E., Altman, D.G., Egger, M., Pocock, S.J., Gøtzsche, P.C., Vandenbroucke, J.P., and STROBE Initiative (2008). The Strengthening the Reporting of Observational Studies in Epidemiology (STROBE) statement: guidelines for reporting observational studies. J Clin Epidemiol 61, 344–349. 10.1016/j.jclinepi.2007.11.008.

87. Karlsen, A., Soendenbroe, C., Malmgaard-Clausen, N.M., Wagener, F., Moeller, C.E., Senhaji, Z., Damberg, K., Andersen, J.L., Schjerling, P., Kjaer, M., et al. (2020). Preserved capacity for satellite cell proliferation, regeneration, and hypertrophy in the skeletal muscle of healthy elderly men. FASEB J 34, 6418–6436. 10.1096/fj.202000196R.

88. Soendenbroe, C., Heisterberg, M.F., Schjerling, P., Kjaer, M., Andersen, J.L., and Mackey, A.L. (2022). Human skeletal muscle acetylcholine receptor gene expression in elderly males performing heavy resistance exercise. Am J Physiol Cell Physiol 323, C159–C169. 10.1152/ajpcell.00365.2021.

89. Jones, C.J., Rikli, R.E., and Beam, W.C. (1999). A 30-s chair-stand test as a measure of lower body strength in community-residing older adults. Res Q Exerc Sport 70, 113–119. 10.1080/02701367.1999.10608028.

90. Lipkin, D.P., Scriven, A.J., Crake, T., and Poole-Wilson, P.A. (1986). Six minute walking test for assessing exercise capacity in chronic heart failure. Br Med J (Clin Res Ed) 292, 653–655. 10.1136/bmj.292.6521.653.

91. Podsiadlo, D., and Richardson, S. (1991). The timed “Up & Go”: a test of basic functional mobility for frail elderly persons. J Am Geriatr Soc 39, 142–148. 10.1111/j.1532-5415.1991.tb01616.x.

92. Bergstrom, J. (1975). Percutaneous needle biopsy of skeletal muscle in physiological and clinical research. Scand. J. Clin. Lab. Invest. 35, 609–616.

93. Fleming, S.J., Chaffin, M.D., Arduini, A., Akkad, A.-D., Banks, E., Marioni, J.C., Philippakis, A.A., Ellinor, P.T., and Babadi, M. (2023). Unsupervised removal of systematic background noise from droplet-based single-cell experiments using CellBender. Nat Methods 20, 1323–1335. 10.1038/s41592-023-01943-7.

94. Wolock, S.L., Lopez, R., and Klein, A.M. (2019). Scrublet: Computational Identification of Cell Doublets in Single-Cell Transcriptomic Data. Cell Syst 8, 281–291.e9. 10.1016/j.cels.2018.11.005.

95. Heaton, H., Talman, A.M., Knights, A., Imaz, M., Gaffney, D.J., Durbin, R., Hemberg, M., and Lawniczak, M.K.N. (2020). Souporcell: robust clustering of single-cell RNA-seq data by genotype without reference genotypes. Nat Methods 17, 615–620. 10.1038/s41592-020-0820-1.

96. 1000 Genomes Project Consortium, Auton, A., Brooks, L.D., Durbin, R.M., Garrison, E.P., Kang, H.M., Korbel, J.O., Marchini, J.L., McCarthy, S., McVean, G.A., et al. (2015). A global reference for human genetic variation. Nature 526, 68–74. 10.1038/nature15393.

97. Hao, Y., Stuart, T., Kowalski, M.H., Choudhary, S., Hoffman, P., Hartman, A., Srivastava, A., Molla, G., Madad, S., Fernandez-Granda, C., et al. (2024). Dictionary learning for integrative, multimodal and scalable single-cell analysis. Nat Biotechnol 42, 293–304. 10.1038/s41587-023-01767-y.

98. Love, M.I., Huber, W., and Anders, S. (2014). Moderated estimation of fold change and dispersion for RNA-seq data with DESeq2. Genome Biol 15, 550. 10.1186/s13059-014-0550-8.

99. Jennekens, F.G., Tomlinson, B.E., and Walton, J.N. (1971). Data on the distribution of fibre types in five human limb muscles. An autopsy study. J. Neurol. Sci. 14, 245–257. 10.1016/0022-510x(71)90215-2.

