## Supplemental Figures and Tables for "Single-Nucleus to Whole Body Phenotyping Reveals Neuromuscular Impairment and Preserved Exercise Adaptations in Long-Term Pediatric HSCT Survivors >10 years after treatment"

This file includes Tables S1–S6 and Figures S1–S7.

**Table S1. Participant characteristics for each part of the study.**

**A** **Baseline**

| | Control (n=28, 64% female) | | | | | | HSCT (n=18, 67% female) | | | | | |
|---|---|---|---|---|---|---|---|---|---|---|---|---|
| | Mean | ± | Std | Min | - | Max | Mean | ± | Std | Min | - | Max |
| Age (yr) | 29 | ± | 7 | 19 | - | 52 | 28 | ± | 6 | 21 | - | 44 |
| Height (cm) | 173 | ± | 8 | 159 | - | 187 | 167 | ± | 11 | 148 | - | 182 |
| Weight (kg) | 69 | ± | 12 | 49 | - | 97 | 61 | ± | 12 | 37 | - | 81 |
| BMI (kg/m$^2$) | 23 | ± | 3 | 18 | - | 28 | 22 | ± | 4 | 16 | - | 30 |

**B** **snRNAseq**

| | Control (n=4, 100% female) | | | | | | HSCT (n=4, 100% female) | | | | | |
|---|---|---|---|---|---|---|---|---|---|---|---|---|
| | Mean | ± | Std | Min | - | Max | Mean | ± | Std | Min | - | Max |
| Age (yr) | 30 | ± | 5 | 24 | - | 35 | 30 | ± | 5 | 23 | - | 33 |
| Height (cm) | 168 | ± | 9 | 159 | - | 179 | 160 | ± | 7 | 152 | - | 168 |
| Weight (kg) | 62 | ± | 10 | 49 | - | 70 | 55 | ± | 15 | 37 | - | 74 |
| BMI (kg/m$^2$) | 22 | ± | 4 | 18 | - | 27 | 22 | ± | 6 | 16 | - | 30 |

**C** **Single Exercise bout**

| | Control (n=11, 100% female) | | | | | | HSCT (n=7, 100% female) | | | | | |
|---|---|---|---|---|---|---|---|---|---|---|---|---|
| | Mean | ± | Std | Min | - | Max | Mean | ± | Std | Min | - | Max |
| Age (yr) | 30 | ± | 9 | 19 | - | 52 | 29 | ± | 5 | 21 | - | 34 |
| Height (cm) | 169 | ± | 7 | 159 | - | 179 | 163 | ± | 8 | 152 | - | 176 |
| Weight (kg) | 67 | ± | 11 | 49 | - | 83 | 62 | ± | 15 | 37 | - | 81 |
| BMI (kg/m$^2$) | 23 | ± | 4 | 18 | - | 28 | 23 | ± | 5 | 16 | - | 30 |

**D** **12-week HReT**

| | Control (n=16, 44% female) | | | | | | HSCT (n=7, 57% female) | | | | | |
|---|---|---|---|---|---|---|---|---|---|---|---|---|
| | Mean | ± | Std | Min | - | Max | Mean | ± | Std | Min | - | Max |
| Age (yr) | 29 | ± | 7 | 22 | - | 49 | 29 | ± | 8 | 22 | - | 44 |
| Height (cm) | 175 | ± | 8 | 160 | - | 187 | 167 | ± | 10 | 152 | - | 177 |
| Weight (kg) | 70 | ± | 13 | 54 | - | 97 | 59 | ± | 10 | 46 | - | 72 |
| BMI (kg/m$^2$) | 23 | ± | 3 | 19 | - | 28 | 21 | ± | 2 | 19 | - | 23 |

Participant characteristics are shown for controls and HSCT survivors included in the full baseline cohort (A), the snRNA-seq analysis (B), the single exercise bout sub-study (C), and the 12-week heavy resistance training (HReT) intervention (D). Data are presented as mean ± SD and minimum–maximum.

**Table S2. Transplant characteristics of HSCT survivors across groups.**

| | All (n=18) | Single exercise (n=7) | 12-week HReT (n=11) |
|---|---|---|---|
| Age at HSCT (yr), median (range) | 11 (1-17) | 10 (6-14) | 12 (1-17) |
| Years from HSCT to study, median (range) | 16 (10-29) | 21 (14-26) | 13 (10-29) |
| **Diagnosis** | | | |
| Malignant diagnosis incl. MDS, n (%) | 16 (89%) | 7 (100%) | 9 (82%) |
| Benign diagnosis, n (%) | 2 (11%) | 0 (0%) | 2 (18%) |
| **Transplantation characteristics** | | | |
| Donor match | | | |
| Matched sibling donor, n (%) | 10 (56%) | 4 (57%) | 6 (55%) |
| Matched unrelated donor, n (%) | 7 (39%) | 3 (43%) | 4 (36%) |
| Other donor, n (%) | 1 (5%) | 0 (0%) | 1 (9%) |
| **Conditioning regimen** | | | |
| Chemotherapy only, n (%) | 8 (44%) | 3 (43%) | 5 (45%) |
| TBI 12GY + CY/VP16 | 10 (56%) | 4 (57%) | 6 (55%) |
| **Acute GvHD** | | | |
| Grade 0-1, n (%) | 9 (50%) | 3 (43%) | 6 (55%) |
| Grade 2-4, n (%) | 9 (50%) | 4 (57%) | 5 (45%) |

Transplant characteristics of HSCT survivors included in the full cohort, the single exercise bout sub-study, and the 12-week heavy resistance training (HReT) trial. Continuous variables are presented as median (range), and categorical variables as n (%).

### Table S3. Blood panel with between-group p values.

| | | | Control | | | | | HSCT | | | | |
|---|---|---|---|---|---|---|---|---|---|---|---|---|
| **Hematology** | Unit | p-value | Mean | Std | Min | Max | n | Mean | Std | Min | Max | n |
| Erythrocytes | ×10¹²/L | 0.54 | 4.49 | 0.30 | 4.04 | 4.99 | 11 | 4.40 | 0.26 | 4.20 | 4.91 | 7 |
| Hemoglobin | mmol/L | 0.84 | 8.7 | 0.8 | 7.3 | 10.0 | 17 | 8.8 | 0.8 | 7.4 | 10.4 | 11 |
| Hematocrit | % | 0.97 | 43 | 3 | 38 | 47 | 17 | 43 | 3 | 36 | 48 | 11 |
| Mean corpuscular volume | fL | 0.62 | 91 | 4 | 86 | 98 | 17 | 92 | 4 | 87 | 97 | 11 |
| Reticulocytes | ×10⁹/L | 0.15 | 64 | 20 | 33 | 101 | 17 | 78 | 27 | 36 | 112 | 11 |
| Platelets | ×10⁹/L | 0.87 | 262 | 61 | 170 | 420 | 28 | 259 | 63 | 158 | 374 | 18 |
| Leukocytes | ×10⁹/L | **0.05** | 6.2 | 1.6 | 3.0 | 8.9 | 28 | 7.5 | 2.7 | 4.2 | 12.7 | 18 |
| Neutrophils | ×10⁹/L | 0.14 | 3.36 | 1.20 | 1.09 | 6.01 | 28 | 4.08 | 2.07 | 1.92 | 9.46 | 18 |
| Lymphocytes | ×10⁹/L | 0.06 | 2.11 | 0.55 | 1.22 | 3.97 | 28 | 2.57 | 1.06 | 1.14 | 4.95 | 18 |
| Monocytes | ×10⁹/L | **0.01** | 0.50 | 0.12 | 0.33 | 0.67 | 11 | 0.79 | 0.31 | 0.47 | 1.23 | 7 |
| Eosinophils | ×10⁹/L | 0.62 | 0.17 | 0.18 | 0.03 | 0.99 | 28 | 0.14 | 0.07 | 0.05 | 0.35 | 18 |
| Basophils | ×10⁹/L | 0.24 | 0.04 | 0.02 | 0.01 | 0.10 | 28 | 0.05 | 0.04 | 0.01 | 0.16 | 18 |
| Immature granulocytes | ×10⁹/L | **0.01** | 0.02 | 0.01 | 0.01 | 0.05 | 11 | 0.04 | 0.02 | 0.02 | 0.08 | 7 |
| **Liver function** | | | | | | | | | | | | |
| Alanine aminotransferase | U/L | 0.55 | 24 | 16 | 12 | 79 | 17 | 27 | 13 | 13 | 62 | 11 |
| Alkaline phosphatase | U/L | **0.03** | 65 | 20 | 38 | 128 | 17 | 83 | 21 | 45 | 112 | 11 |
| Bilirubin (total) | µmol/L | 0.72 | 9 | 3 | 4 | 15 | 17 | 9 | 8 | 3 | 27 | 11 |
| **Kidney function and electrolytes** | | | | | | | | | | | | |
| Creatinine | µmol/L | 0.45 | 71 | 12 | 58 | 103 | 17 | 76 | 19 | 50 | 112 | 11 |
| Sodium | mmol/L | 0.15 | 139 | 2 | 132 | 142 | 17 | 141 | 2 | 136 | 143 | 11 |
| Potassium | mmol/L | 0.34 | 3.7 | 0.2 | 3.3 | 4.2 | 16 | 3.8 | 0.3 | 3.4 | 4.1 | 11 |
| Urine albumin | mg/L | 0.28 | 39 | 67 | 3 | 280 | 17 | 80 | 123 | 5 | 373 | 9 |
| **Thyroid function** | | | | | | | | | | | | |
| Thyroid-stimulating hormone | mU/L | 0.09 | 1.96 | 0.85 | 0.71 | 4.06 | 17 | 2.86 | 1.84 | 0.92 | 6.30 | 11 |
| Free thyroxine | pmol/L | 0.23 | 16.3 | 2.3 | 12.2 | 20.9 | 17 | 24.2 | 26.3 | 12.8 | 103.0 | 11 |
| Triiodothyronine | nmol/L | **0.01** | 1.64 | 0.21 | 1.20 | 1.90 | 17 | 1.95 | 0.34 | 1.40 | 2.50 | 11 |
| **Lipid profile** | | | | | | | | | | | | |
| Total cholesterol | mmol/L | 0.67 | 4.3 | 0.9 | 3.0 | 5.9 | 17 | 4.5 | 0.8 | 3.4 | 6.3 | 11 |
| HDL cholesterol | mmol/L | 0.23 | 1.5 | 0.4 | 0.9 | 2.4 | 17 | 1.3 | 0.3 | 0.7 | 1.9 | 11 |
| LDL cholesterol | mmol/L | 1.00 | 2.4 | 0.8 | 1.2 | 3.7 | 17 | 2.4 | 0.5 | 1.7 | 3.3 | 10 |
| Triglycerides | mmol/L | 0.09 | 1.0 | 0.5 | 0.3 | 2.2 | 17 | 1.6 | 1.3 | 0.4 | 4.5 | 11 |
| **Miscellaneous** | | | | | | | | | | | | |
| HbA1c | mmol/L | 0.08 | 5.7 | 0.5 | 4.6 | 6.5 | 11 | 6.2 | 0.6 | 5.2 | 6.8 | 7 |
| Ferritin | µg/L | **0.03** | 105 | 94 | 17 | 307 | 16 | 198 | 121 | 36 | 401 | 11 |
| 25-hydroxyvitamin D | nmol/L | **0.03** | 48 | 21 | 11 | 108 | 17 | 69 | 26 | 19 | 117 | 11 |
| Immunoglobulin G | g/L | 0.13 | 9.9 | 2.2 | 6.3 | 14.4 | 17 | 8.7 | 1.4 | 6.3 | 10.8 | 11 |
| C-reactive protein | mg/L | 0.46 | 6 | 7 | 1 | 21 | 8 | 8 | 9 | 1 | 34 | 13 |
| Creatine kinase | U/L | 0.16 | 72 | 33 | 38 | 143 | 11 | 100 | 49 | 36 | 184 | 7 |
| Haptoglobin | g/L | 0.15 | 1.05 | 0.38 | 0.38 | 1.69 | 16 | 1.30 | 0.52 | 0.46 | 1.94 | 11 |

Hematological and biochemical blood variables measured in controls and HSCT survivors. For each variable, the table shows the measurement unit, between-group p value, and descriptive statistics (mean, SD, minimum, maximum, and n) for the Control and HSCT groups.

**Table S4. Twelve-week heavy resistance training program.**

| Session # | 1-3 | 4-9 | 10-16 | 17-23 | 24-30 | 31-36 |
|---|---|---|---|---|---|---|
| Leg press | | | | | | |
| % 1RM | 60 | 69 | 74 | 77 | 79 | 84 |
| Sets per session | 3 | 4 | 5 | 5 | 4 | 4 |
| Repetitions per set | 12 | 10 | 10,10,10,8,8 | 10,10,8,8,6 | 8,8,6,6 | 8,6,6,4 |
| Knee extension | | | | | | |
| % 1RM | 60 | 69 | 74 | 77 | 79 | 79 |
| Sets per session | 3 | 4 | 4 | 5 | 5 | 4 |
| Repetitions per set | 12 | 10 | 10 | 10,10,8,8,8 | 8,8,8,6,6 | 8,8,6,6 |
| Hamstring curl | | | | | | |
| % 1RM | 60 | 69 | 74 | 77 | 79 | 79 |
| Sets per session | 3 | 4 | 4 | 4 | 4 | 4 |
| Repetitions per set | 12 | 10 | 10 | 10,10,8,8 | 8 | 8,8,6,6 |

Progressive heavy resistance training (HReT) program performed over 36 supervised sessions. For each exercise, the table shows the prescribed relative load (%1RM), number of sets per session, and repetitions per set across the intervention.

**Table S5. Primers used for qPCR analyses.**

| mRNA | Genbank ID | Sense | Antisense |
|---|---|---|---|
| **HUMAN** | | | |
| RPLP0 | NM_001002 | GGAAACTCTGCATTCTCGCTTCCT | CCAGGACTCGTTTGTACCCGTTG |
| XRRA1 Exon 1-2 | NM_182969 | CAGACCCCTCGGAGGCTGAA | TGGGAATGGCCCCTTAACTTCC |
| XRRA1 Exon 2-3 | NM_182969 | AGGGGCCATTCCCAAAGTCAA | CAGGTAAGGCTTCCCATCATCCA |
| XRRA1 Exon 2-4 | NM_001378162 | AGGGGCCATTCCCAAAGTCAA | CCCTTGGGCTTCTTCTTGAGGT |
| XRRA1 Exon 2-4 (no 3) | NM_001378162 | AGGGGCCATTCCCAAAGTCAA | CCACTAACCAGTGTCCTTGGCTTG |
| **RAT** | | | |
| Rplp0 | NM_022402 | CCAGAGGTGCTGGACATCACAGAG | TGGAGTGAGGCACTGAGGCAAC |
| Gapdh | NM_001289726 | CAGCAACTCCCACTCTTCCACCT | ACCACCCTGTTGCTGTAGCCGT |
| Fth1 | NM_012848 | GCACTGCACTTGGAAAAGAGTGTGAA | CCTGCTCATTCAGGTAATGCGTCT |
| Ppp1r1a | NM_022676 | AGACACAGGCTCAGCGTCAAGG | TGCTCCTGAGTCTTGGGTTTGG |
| Rack1 | NM_130734 | GCCACCCCAGTGTACCTCTTTG | TCACCTGCCATACACGCACCAA |
| Vimentin | NM_031140 | TCCTCTGGTTGACACCCACTCC | GTTTTTATTCAAGGTCATCGTGGTGCT |
| Cdh13 | NM_138889 | GCCTCAGCTTGCTGCTGCTCT | GGGAGTCAAGCTTCAGATGTGTCGT |

Primer sequences used for qPCR analyses in human and rat samples, including target mRNA, GenBank accession number, and sense and antisense sequences.

**Table S6. Primary antibodies and dyes used in immunofluorescence staining protocols.**

| PRIMARY ANTIBODIES | | | | | |
|---|---|---|---|---|---|
| **Host** | **Antibody** | **Company** | **Cat. no.** | **Concentration** | **RRID** |
| Rabbit | NCAM | Sigma-Aldrich | AB5032 | 1:250 | AB_11213653 |
| Mouse, IgG1 | MyHC I | DSHB | A4.951 | 1:100 | AB_10540570 |
| Mouse, IgG2b | Dystrophin | Sigma-Aldrich | D8168 | 1:500 | AB_259245 |
| Mouse, IgG2b | MyHC I | DSHB | BA.D5 | 1:100 | AB_2235587 |
| Rabbit | PCM1 | Sigma-Aldrich | HPA023370 | 1:1000 | AB_1855072 |
| Goat | Collagen IV | Millipore | AB769 | 1:500 | AB_92262 |
| Rabbit | Laminin | Sigma-Aldrich | L9393 | 1:200 | AB_477163 |
| Rabbit | Desmin | Abcam | AB32362 | 1:1000 | AB_731901 |
| Mouse, IgG1 | Myogenin | DSHB | F5D | 1:50 | AB_2146602 |
| Rabbit | XRRA1 | Thermo Fisher Scientific | PA5-65227 | 1:100 | AB_2663700 |
| Rabbit | Tau1 | GeneTex | GTX130462 | 1:500 | AB_2886280 |
| Goat | ChAT | Millipore | AB144p | 1:100 | AB_11212924 |
| Mouse, IgG2b | Ki67 | Abcam | ab238020 | 1:1000 | AB_3076661 |

| SECONDARY ANTIBODIES | | | | | |
|---|---|---|---|---|---|
| **Host** | **Antibody** | **Company** | **Cat. no.** | **Concentration** | **RRID** |
| Goat | Anti-rabbit, 488 | Invitrogen | A-11034 | 1:200 | AB_2576217 |
| Goat | Anti-mouse IgG2b, 568 | Invitrogen | A-21144 | 1:500 | AB_2535780 |
| Goat | Anti-mouse IgG1, 647 | Invitrogen | A-21240 | 1:500 | AB_2535809 |
| Donkey | Anti-mouse, 488 | Abcam | ab150109 | 1:200 | AB_2571721 |
| Donkey | Anti-rabbit, 568 | Abcam | ab175693 | 1:200 | AB_2884939 |
| Donkey | Anti-goat, 680 | Invitrogen | A-21084 | 1:200 | AB_141494 |
| Goat | Anti-mouse, IgG2b, 488 | Invitrogen | A-21141 | 1:500 | AB_141626 |
| Goat | Anti-rabbit, 680 | Invitrogen | A-21076 | 1:500 | AB_2535736 |

| DYES | | | | | |
|---|---|---|---|---|---|
| **Host** | **Dye** | **Company** | **Cat. no.** | **Concentration** | **RRID** |
| N/A | Wheat Germ Agglutinin | Invitrogen | W32465 | 1:200 | N/A |
| N/A | Hoechst | Invitrogen | H1399 | 1:200 | N/A |
| N/A | DAPI | Thermo Fisher Scientific | P36931 | N/A | N/A |

Overview of primary antibodies, secondary antibodies, and dyes used for immunofluorescence staining. For each reagent, the table shows host species, antigen or reagent name, supplier, catalog number, working concentration, and RRID, where available.

**Figure S1. Glucose and C-peptide responses during a 3-hour oral glucose tolerance test.**

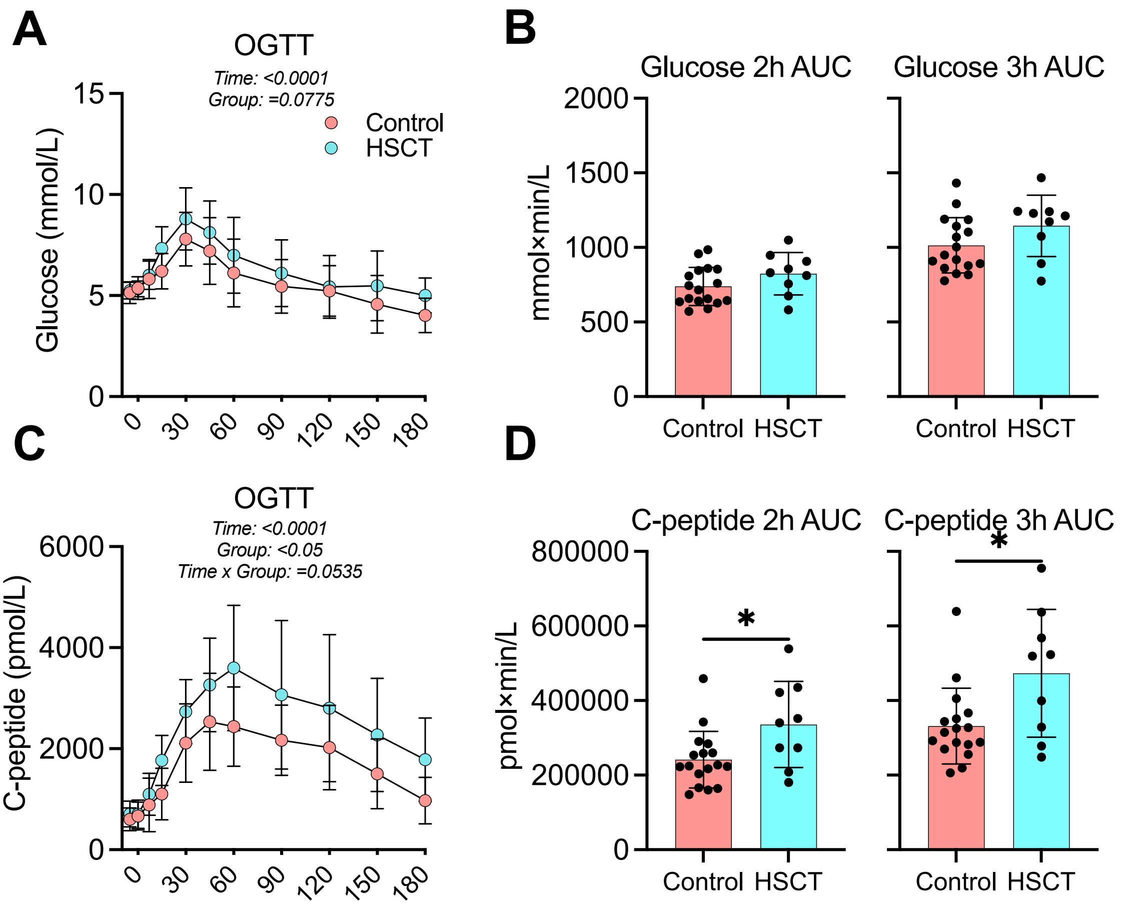

(A) Plasma glucose concentrations during a 3-h oral glucose tolerance test (OGTT) at baseline. (B) Glucose area under the curve (AUC) calculated for the first 2 h and the full 3 h of the OGTT. (C) Plasma C-peptide concentrations during the 3-h OGTT. (D) C-peptide AUC calculated for the first 2 h and the full 3 h of the OGTT. AUC comparisons were analyzed using unpaired two-tailed t tests. *p < 0.05.

**Figure S2. PBMC single-nucleus RNA sequencing analysis in HSCT survivors and healthy controls.**

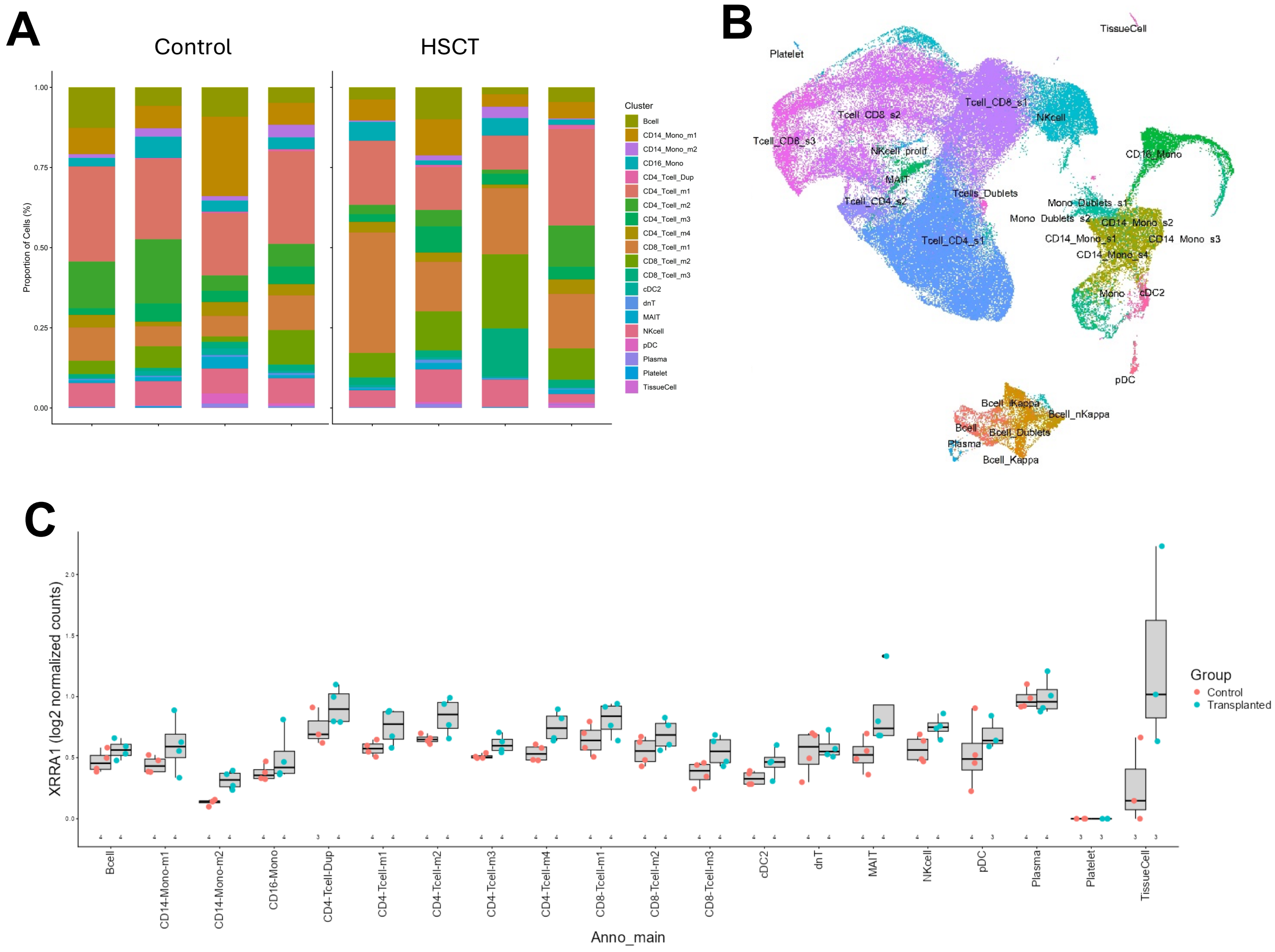

Single-nucleus RNA sequencing (snRNA-seq) was performed on peripheral blood mononuclear cells (PBMCs) from four HSCT survivors and four matched healthy controls. (A) Proportion of nuclei assigned to each major immune cell cluster across groups. (B) UMAP of subclusters. (C) XRRA1 expression across the main PBMC clusters.

**Figure S3. Skeletal muscle single-nucleus RNA sequencing in HSCT survivors and healthy controls.**

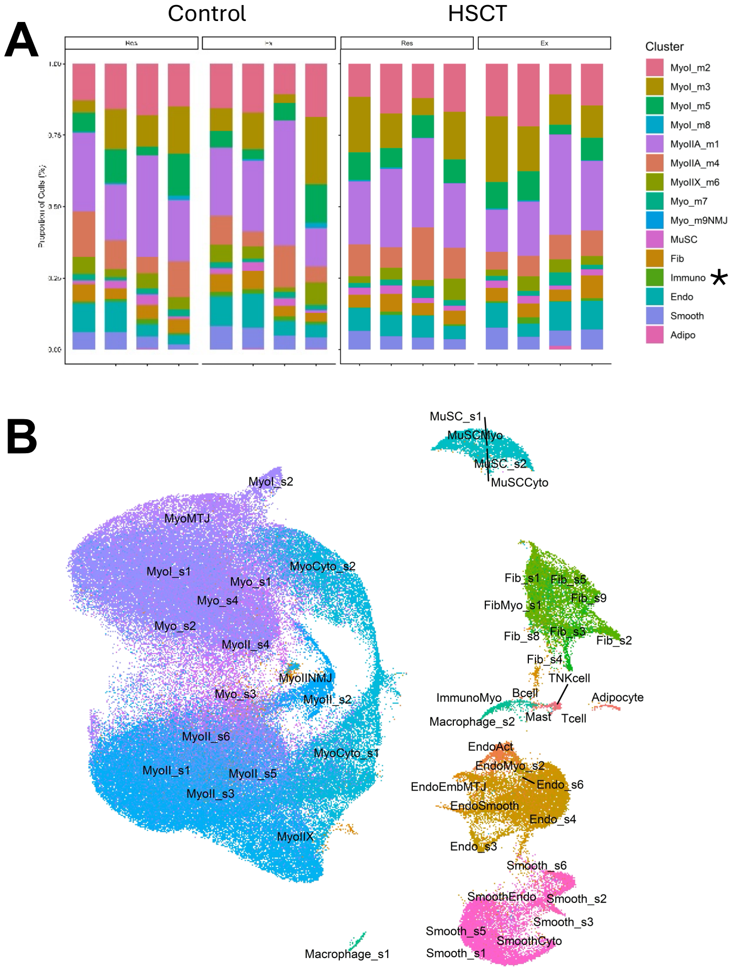

Single-nucleus RNA sequencing (snRNA-seq) was performed on skeletal muscle biopsies from four HSCT survivors and four matched healthy controls, from both the exercised and contralateral leg 7 days post exercise (16 samples in total). (A) Proportion of nuclei assigned to each major muscle-resident cell population across groups; * indicates significant differences between legs (effect of exercise). (B) UMAP of subclusters.

**Figure S4. Representative XRRA1 immunofluorescence staining in skeletal muscle.**

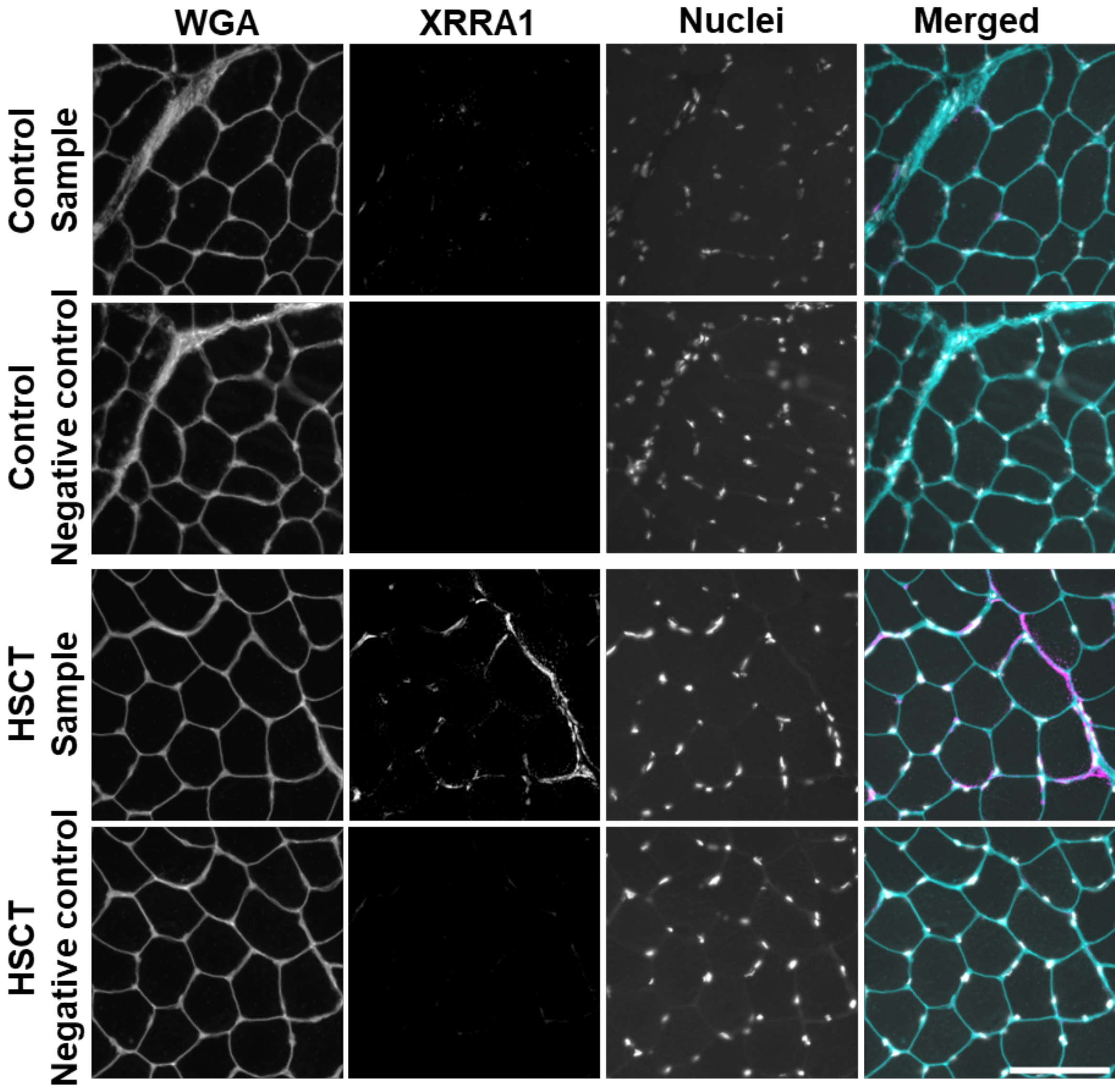

Representative images of XRRA1 immunofluorescence in muscle sections from a control participant (two top rows) and an HSCT survivor (two bottom rows). For each sample, images obtained with and without primary antibody incubation are shown to illustrate background signal. Scale bar, 100 µm.

**Figure S5. Exercise-associated transcriptional responses across skeletal muscle cell populations.**

### Muscle snRNAseq exercise DEGs

**A**

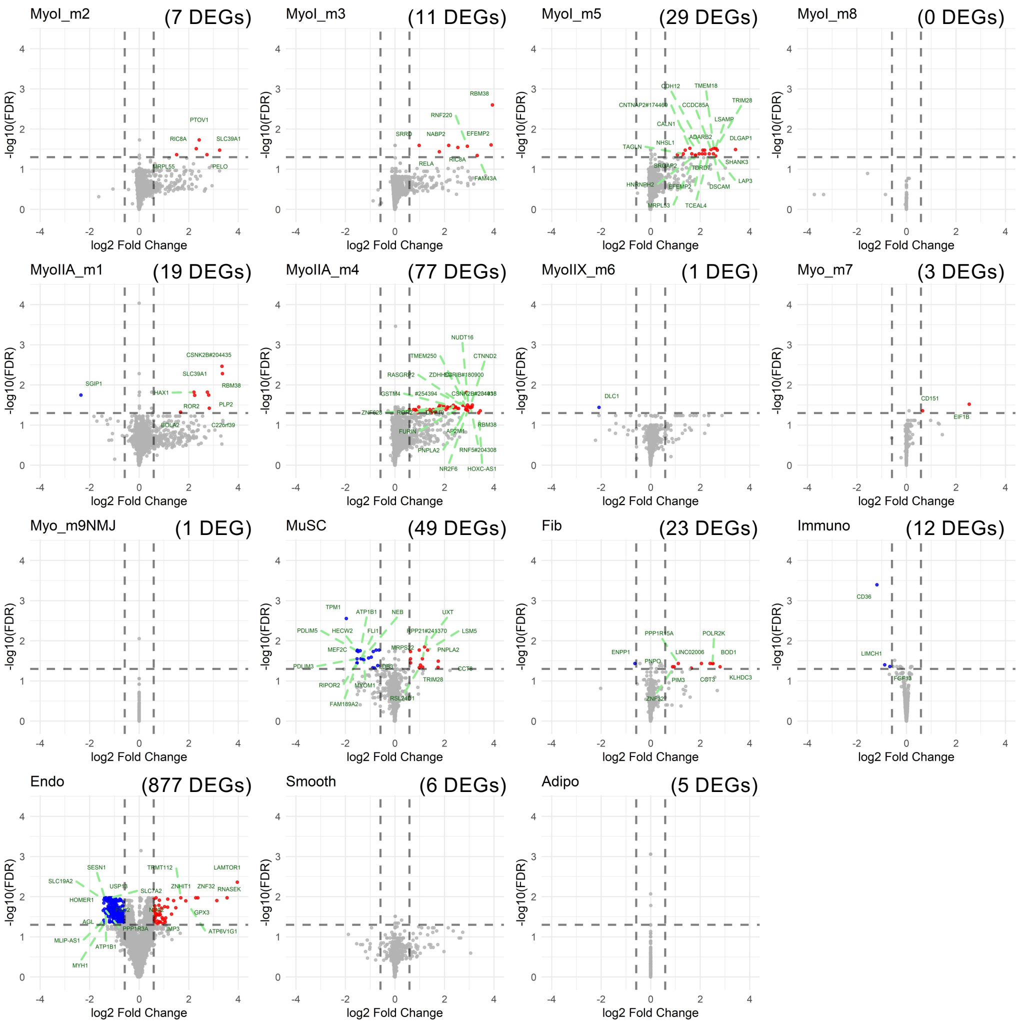

**B**

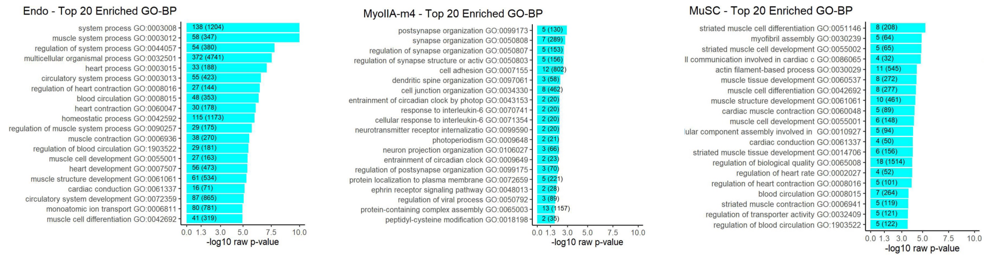

Single-nucleus RNA sequencing (snRNA-seq) was used to assess the 7-day transcriptional response to exercise in human skeletal muscle. (A) Main effect of exercise (difference between legs) shown as the number of differentially expressed genes (DEGs) identified within each main cell cluster. (B) Top 20 GO-BP pathways enriched among DEGs in the three cell types with the largest number of exercise-responsive genes: endothelial cells, myonuclei (MyoIIA-m4), and muscle stem cells (MuSCs).

**Figure S6. Changes in maximal strength during the 12-week resistance training intervention.**

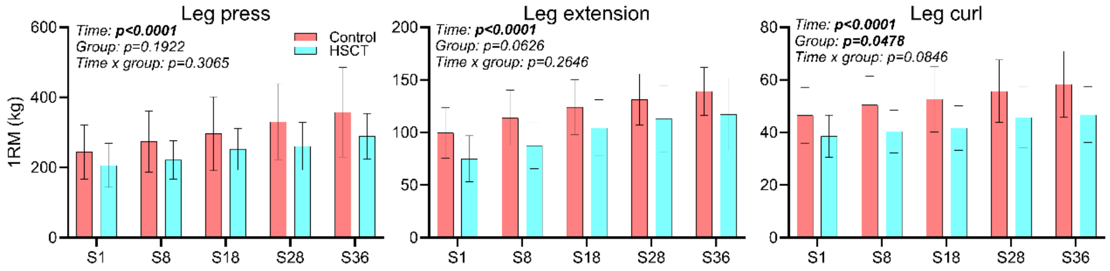

One-repetition maximum (1RM) changes in leg press, leg extension, and leg curl over the course of the 12-week training intervention in controls (n = 15) and HSCT survivors (n = 7). Data were analyzed using a two-way mixed-effects model (group × time). Values are shown as mean ± SD.

**Figure S7. Flow chart (12-week training intervention).**

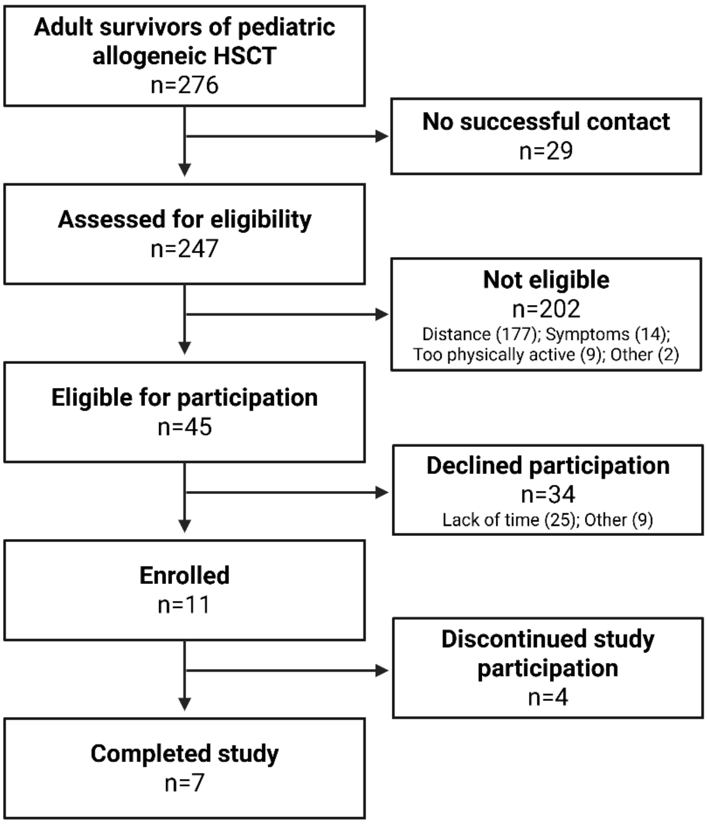

Flow chart for HSCT survivors in 12-week training intervention
